# Care-cascade failures as a determinant of infectious burden in tuberculosis: an agent-based modelling study in India

**DOI:** 10.64898/2026.10.27.26366617

**Authors:** Youngji Jo, Jonathan Bloedow, Sourya Shrestha, Edward Wenger, Katia Bulekova, Stewart Chang

## Abstract

**Background:** Despite effective treatment, care failures can prolong tuberculosis (TB) infectiousness, sustain transmission and hinder progress towards End TB targets. Counts of patients diagnosed and treated obscure these delays. We examined how care-cascade failures shape infectious burden and intervention impact in India.

**Methods:** We used an existing EMOD-TBHIV agent-based modelling platform for India, calibrated the model to national incidence, prevalence and mortality, and assessed agreement with published care-cascade benchmarks. We classified simulated TB disease episodes by diagnostic failures (false-negative tests or pre-treatment default), treatment failure, both, or neither. We quantified infectiousness-weighted person-time across presymptomatic disease, patient delay and health-system delay. We projected effects of active case finding, improved diagnostic sensitivity and treatment support during 2025–35, individually and combined, and examined sensitivity across 20 calibrated parameter sets.

**Findings:** Among 25,894 simulated TB episodes beginning in 2015–24, 39% had neither recorded failure type, 26% had diagnostic failures only and 35% had treatment failure, with or without diagnostic failures. Median effective infectious time was 3 months among episodes with neither failure type versus 16 months among those with treatment failure. The latter accounted for 63% of infectiousness-weighted burden. Overall, 64% of active TB person-time accrued after the first diagnostic test. Treatment support and active case finding reduced 2035 incidence relative to baseline by 18% (95% CI 15–21) and 17% (13–20), respectively; Diagnostic improvement reduced incidence by 3% (−1 to 6), and the combined package by 34% (31–36). Relative to 2015, the package reduced prevalence by 85% and deaths by 89% but incidence by only 57%, short of the 90% End TB target in every parameter set.

**Interpretation:** Repeated care failures prolonged infectiousness among patients already in care, so that supporting treatment completion reduced transmission as much as finding new cases. Monitoring their frequency and duration alongside conventional cascade indicators could identify opportunities to shorten infectious time and inform programme priorities.

## Introduction

Despite being curable in the vast majority of cases, tuberculosis (TB) is the leading cause of infectious mortality worldwide, causing over 1.25 million deaths in 2023[1]. A major contributor is underdiagnosis and undertreatment: prevalence surveys in Asia consistently find that half or more of people with prevalent, undiagnosed TB have already sought care at a medical facility[2], and a systematic review estimated that 13% of people diagnosed with bacteriologically confirmed TB in high-burden countries were lost to follow-up before starting treatment[3]. People who have TB, seek care for their symptoms and still do not receive appropriate diagnosis and treatment represent a large missed opportunity to reduce transmission and mortality. India accounts for a quarter of the global burden, the highest of any country, and its fragmented public and private sectors mean patients often visit a series of providers before being diagnosed and treated[4]. The WHO End TB Strategy targets a 90% reduction in TB incidence and a 95% reduction in TB deaths by 2035 relative to 2015[5, 6]; by 2021 global incidence had fallen only about 10% and deaths 5.9%, far short of the 2020 milestones of 20% and 35%.[7] The Lancet Commission identified insufficient case finding and diagnosis as the central challenge, with only 38% of notified patients receiving a WHO-recommended rapid molecular test at diagnosis in 2021.[8]

Despite three decades of DOTS expansion and the post-2012 GeneXpert rollout, India’s incidence has declined more slowly than the End-TB target requires[9]. The government has set a goal of raising early case finding from 14% to 75%, largely through engagement of private providers[10], who treat more than half of estimated cases and are often the first point of contact even for patients later treated in the public sector, but whose diagnostic and treatment practices frequently generate repeat testing and repeat treatment through false negatives and loss to follow-up; patient-level and health-system factors both contribute[11]. These delays worsen individual outcomes and lengthen infectious periods, sustaining transmission at the community level[12]: about half the household contacts of a person with TB in low- and middle-income settings are already infected (51.5%), and their risk of disease is greatest in the first year after exposure[13].

Recent TB modelling has increasingly highlighted the importance of tracking cumulative disease burden beyond conventional programmatic outcomes. TB burden extends past active disease to post-treatment morbidity and mortality, with post-TB sequelae substantially increasing estimated lifetime burden[14–16], and because that downstream risk depends partly on the duration and severity of preceding active disease, individual disease histories matter for quantifying both transmission and longer-term burden[17]. Ku et al.[18]estimated asymptomatic, symptomatic and care-seeking durations by Bayesian calibration, and Ryckman et al.[19] characterised smear- and symptom-specific trajectories, identifying smear-positive subclinical TB as an important transmission reservoir. These approaches focus on natural history and pre-diagnosis trajectories, leaving heterogeneity in post-diagnosis diagnostic and treatment pathways less well characterised.

Agent-based models provide a unique opportunity to move from aggregate cascade outcomes to individual care trajectories by preserving the complete sequence and timing of events experienced by each patient. The EMOD-TBHIV platform[20] used in this study tracks repeated diagnostic attempts, treatment initiation, default, and treatment failure, allowing cumulative infectious person-time to be reconstructed for each TB episode. Yet this longitudinal information has rarely been used to examine how different pathways through the care cascade generate heterogeneity in disease duration and transmission. We leverage this capability to reconstruct patient-level care trajectories, identify recurring patterns of diagnostic and treatment looping, and quantify the disproportionate infectious burden associated with these pathways— burden that is obscured when programs are evaluated only by aggregate cascade indicators.

## Methods

TB transmission dynamics were simulated using EMOD, a software platform for agent/individual-based epidemiological modeling of infectious diseases. EMOD consists of the three frameworks that can define demographics, disease dynamics (configuration), and intervention (campaign) components.

### Population

We configured the EMOD-TBHIV individual-based simulation (DTK v2.21) for India-specific demographics, age-specific contact structure, and heterogeneity in transmission and care quality. Non-disease mortality was applied by age and calendar year from an India-specific all-cause mortality series spanning 1991-2018, with the birth rate scaled so that the simulated population reproduces India’s growth and ageing to 2050; the series is described in Appendix Text S-1. Geography entered as stratified mixing rather than explicit locations: individuals were assigned in equal thirds to low-, medium- or high-burden strata with strongly assortative contact, and age mixing followed the India contact matrix in four groups (0-19, 20-39, 40-59 and >=60 years)[21]. Care quality was a separate individual property (Table 1) governing each individual’s cascade parameters without altering contact. Full parameterisation is given in Appendix Text S-1.

### Natural history of TB

Individuals transitioned among susceptible, latent, active pre-symptomatic, active symptomatic and recovered states (Appendix Figure S-1). After infection they entered fast- or slow-progressing latency, with age-dependent fast progression (3% under 15 years, 7.6% in adults) and a calibrated reactivation rate of 0.30% per year[22]; latent infection was non-infectious and did not self-clear, but remained open to exogenous reinfection, which returned the individual to the fast-progression draw. Active disease began with a pre-symptomatic phase of reduced infectiousness. Symptomatic TB was smear-positive, smear-negative (about 35% as infectious) or extrapulmonary (non-infectious), and without treatment could resolve spontaneously or end in TB-related death. Previously infected individuals, whether latent or recovered after treatment, had about 68% reduced susceptibility to reinfection, with relapse tracked separately from durable cure. Natural-history parameters were held constant throughout and are listed in Appendix Text S-1 and Table S-9.

### Care access and historical interventions

Symptomatic individuals sought care through higher-or lower-quality pathways differing in diagnostic performance and in treatment outcomes; time to care seeking was represented by fast- and slow-seeking subgroups common to both. The model incorporated DOTS expansion from 1997 and GeneXpert scale-up during 2012-2020, with testing and treatment outcomes specific to setting and period (Appendix Text S-1). Beyond these dated milestones, three gradual trends were fitted rather than assumed, because together they determine how much of India’s observed incidence decline the care cascade is credited with. Two act inside the cascade: the fraction of diagnosed patients who initiate treatment rises from 0.70 in 1997 to 0.75 in 2024 in both streams, and the per-timestep probability of seeking care rises geometrically, shortening the mean time to first contact 1.6-fold over the same period. The third acts outside it: a node-level infectivity multiplier scales the force of infection by (1 - d) each year from 1997 to 2024 and then holds it constant, with d calibrated jointly with every other parameter. This term is a deliberately agnostic stand-in for determinants of transmission the cascade does not represent - household crowding and housing quality, nutrition, indoor air pollution, and the age-structure shift of the Indian population. Because d = 0 lies inside its prior range, the calibration was free to attribute the entire observed decline to cascade improvement; it did not, returning d = 0.0128 per year, a cumulative 0.71-fold reduction in non-cascade transmission by 2024 (Table 1). All three trends are frozen at their 2024 values from 2025 onward, so the projected baseline declines on epidemic momentum and interventions introduced from 2025 are evaluated as additional to it.

**Table 1.** Baseline model parameters and assumptions for tuberculosis transmission and care cascades.

| Category | Parameter | Value | Uncertainty range | Source |
| --- | --- | --- | --- | --- |
| **Natural History of Disease** | Base infectivity | 0.0420 | 0.005-0.1 | Calibrated (this study) |
|  | Mean active TB duration (untreated) | 3.5 yr | Fixed | Tiemersma et al. 2011[23]. Nominal input; together with the case-fatality ratio and a 1.5x multiplier on the derived mortality rate it gives a natural history untreated duration of 2.7 years for smear-positive disease (3.6 years across presentations, absent any care). |
|  | Mean presymptomatic duration | 0.233 yr (~85 d) | 0.15-0.5 | Ku et al. 2021[18]; Emery et al. 2023[24]. Ku et al. did not include India; the closest comparators are Pakistan (4.9 months) and Viet Nam (6.5 months). Ku et al.'s phase ends at the survey symptom threshold, typically two weeks of cough, whereas the model's phase ends at symptom onset (resulting in a shorter mean duration). |
|  | Reactivation rate (slow progressor) | 8.27e-06 | 5e-6 to 5e-5 | Calibrated (this study) |
|  | Re-infection susceptibility | 0.32 | 0.2-0.8 | Prior informed by Andrews 2012[25]; calibrated (this study) |
|  | Fast-progressor fraction (adults) | 0.076 | 0.05-0.2 | Calibrated (this study) |
|  | Subclinical (presymptomatic) infectiousness mult. | 0.166 | Fixed | Calibration design value (this study); cf. Emery et al. 2023[24]. A scale factor on Base_Infectivity, i.e. relative to a smear-positive case. |
|  | Smear-positive fraction (adults) | 0.65 | Fixed | Model assumption |
|  | Smear-negative infectiousness multiplier | 0.346 | Fixed | Behr et al. 1999[26] |
|  | Untreated case-fatality ratio | 0.55 | Fixed | Calibration design value (this study). Tiemersma et al. 2011[23] report ~70% 10-year case fatality for untreated smear-positive disease; 0.55 is the nominal input; with the 1.5x mortality multiplier the realised ratio is 0.65 for smear-positive disease and 0.53 across all active TB, which includes smear-negative and extrapulmonary episodes (Appendix Text S-1.2). |
|  | Annual secular decline in transmission outside the cascade, 1997-2024 | 0.0128 /yr (x0.71 by 2024) | 0-0.020 | Calibrated (this study); proxy for living conditions, nutrition, demographic change |
| **Cascade (baseline)** | Population share served by higher- vs lower-quality care streams | 15% / 85% | Fixed | Assumed (the lower-quality stream spans private, informal, and unqualified providers as well as under-resourced public facilities); private-sector share informed by Arinaminpathy et al. 2016[27] |
|  | Care-seeking delay (fast subgroup): Mean days from symptom onset to first care | 14.2 d in 1997, improving to 9.1 d by 2025 | 10-30 | Calibrated (this study) |
|  | Care-seeking delay (slow subgroup): Mean days from symptom onset to first care | 158 d in 1997, improving to 101 d by 2025 | 60-180 | Calibrated (this study) |
|  | GeneXpert sensitivity (SmPos / SmNeg, High QoC) | 0.75 / 0.65 | Fixed | Steingart et al. 2014[28] |
|  | Smear sensitivity (SmPos / SmNeg, Low QoC) | 0.65 / 0.60 | Fixed | Steingart et al. 2006[29] |
|  | Care-seeking budget (Max healthcare visits per TB episode) | 7 visits | Fixed | Model assumption |
|  | Treatment-initiation fraction (both care streams) | 0.70 in 1997, rising to 0.746 by 2024 | Calibrated improvement rate | Calibrated (this study) |
|  | Treatment cure proportion, low- vs high-quality stream | 0.50 / 0.80 | Fixed | emodpy-tbhiv standard drug definitions; Subbaraman 2016[3] |

Note: Across episodes beginning in 2015–2024 the model yields a mean of 1.45 years from activation to resolution (median 1.07): about 3 months presymptomatic, 3 months of patient delay, 2 months untreated after a first test, 5 months in unresolved treatment failure and 4 months on effective treatment — or 0.93 years weighted by relative infectiousness (presymptomatic 23%, untreated 100%, treated 0%).

### Model and cascade-trajectory classification

Each agent progresses through infection, latency and, upon activation, a diagnostic and treatment cascade whose events are logged per episode: infection, pre-symptomatic and symptomatic onset, testing and outcomes, treatment initiation and failure, and resolution by cure or death. To analyze patient trajectories, we classified each active TB episode into one of four mutually exclusive groups based on two types of cascade triggers: (1) **diagnostic loops** (false-negative test results or pre-treatment default) and (2) **treatment loops** (regimen treatment failure requiring re-treatment). Episodes were categorized as **No Loop** (neither diagnostic nor treatment failure occurred, encompassing cases cured on first attempt or never tested), **Diagnostic Loop Only** (experienced diagnostic delay or default, but treatment succeeded once initiated), **Treatment Failure Only** (experienced treatment failure without prior diagnostic delays), or **Diagnostic + Treatment Loop** (experienced both diagnostic delays and treatment failure). For burden analyses, all episodes with treatment failure were consolidated into a single **Any Treatment Failure** stratum (35% of episodes). This produced three primary analytical strata—*No Loop* (39%), *Diagnostic Loop Only* (26%), and *Any Treatment Failure* (35%)—providing a trajectory-based counterpart to aggregate cascade benchmarks (Figure 1A, Table 2).

### Infectious-time decomposition

To quantify each pathway’s transmission contribution, we reconstructed every episode’s infectious period from its event timestamps and partitioned it into three sequential phases (Table 2): the subclinical phase (presymptomatic activation to symptomatic onset), the patient-delay phase (symptomatic onset to first diagnostic test), and the health-system-delay phase (first test to resolution, encompassing all diagnostic and treatment loops). Because infectiousness differs across states, we weighted each phase by its relative transmissibility — ≈23% subclinical, 100% during untreated active disease (patient delay and unresolved loops), and 0% on effective treatment and after cure — and defined the effective infectious time as the infectiousness-weighted sum of phase durations. Figure 1B contrasts three illustrative patient trajectories with progressively greater cumulative infectious burden. Patient A (no loop) has a total episode duration of 8.6 months and accrues approximately 4.4 months of effective infectious time. Patient B (diagnostic loop) accumulates 9.3 months of effective infectious time over 13.5 months, while Patient C (diagnostic loop plus treatment failure) accumulates 22.9 months over 34.1 months. Effective infectious time weights the duration spent in each state by its relative infectiousness. This trajectory-level decomposition captures how diagnostic delays, repeated testing, treatment failure, and re-treatment contribute to transmission burden beyond what aggregate cascade-stage retention alone can reveal. The episode cohort comprises every active TB episode activating in 2015-2024, the decade before the interventions begin, that resolves within the simulation; it describes the contemporary cascade rather than the full 1850-2050 run, more than half of whose episodes predate the DOTS programme.

**Figure 1.**
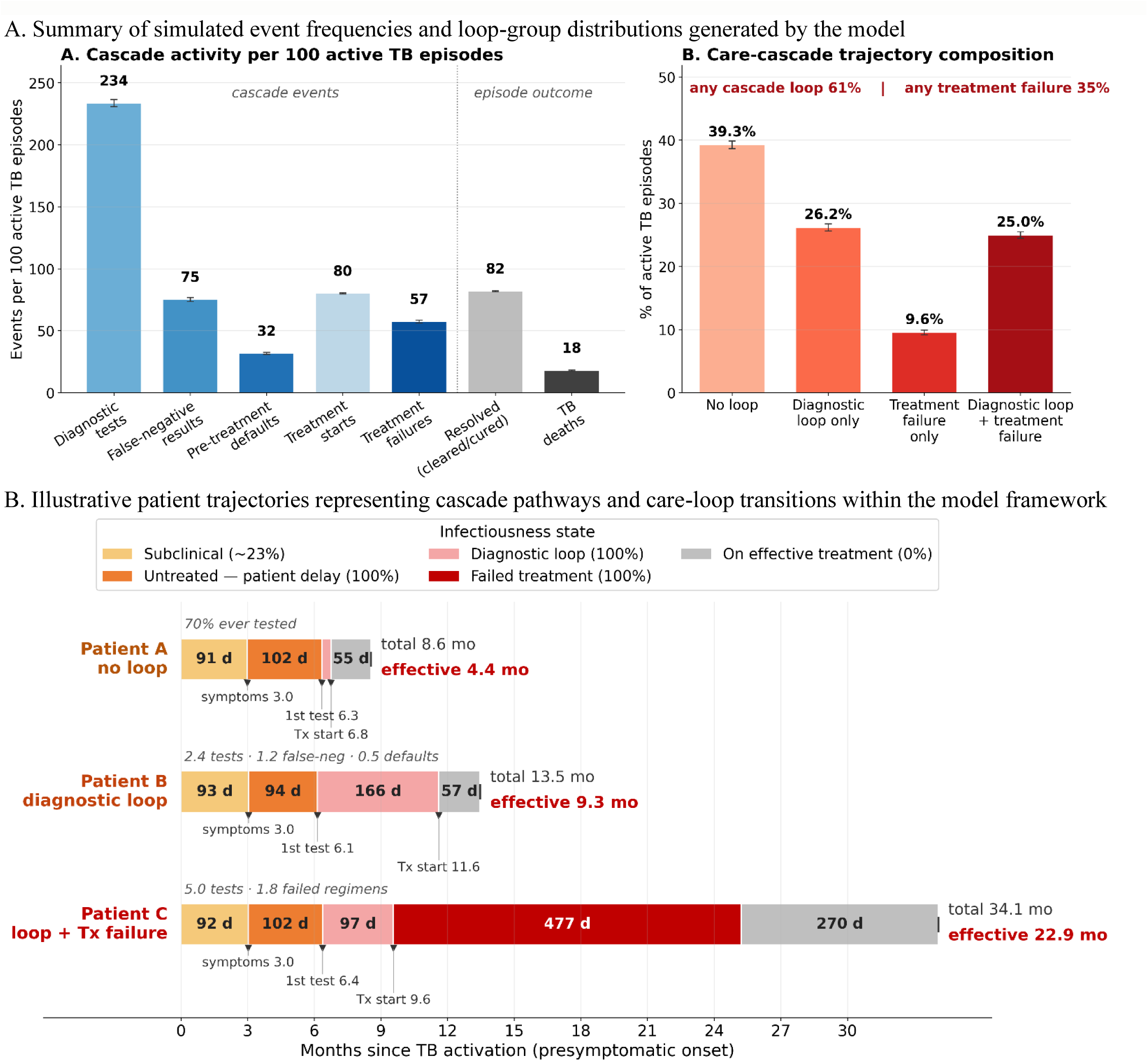
Simulated care-loop dynamics in the agent-based tuberculosis cascade model Note: Baseline scenario, 100 replicates, 25,894 active TB episodes activating in 2015–2024 and followed to resolution.

**Table 2.**
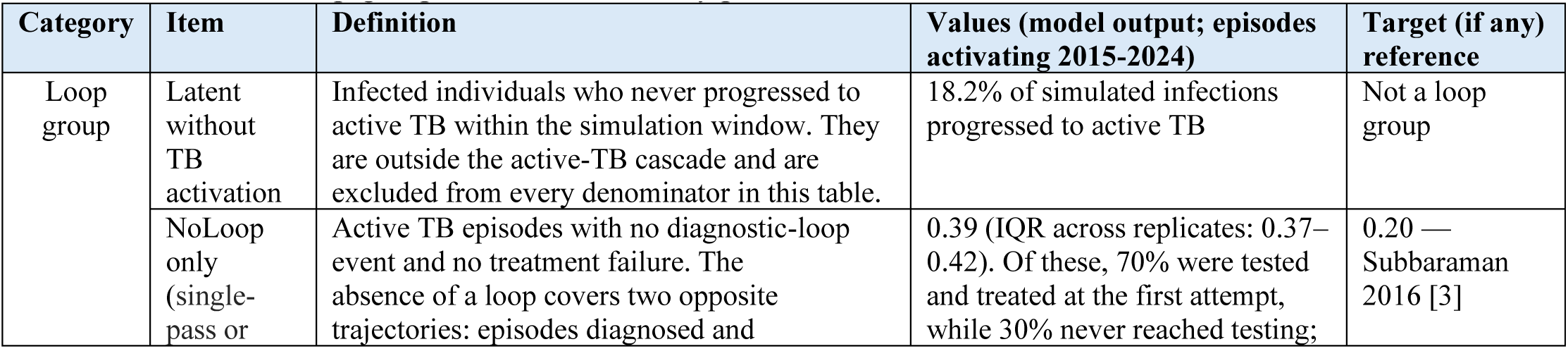
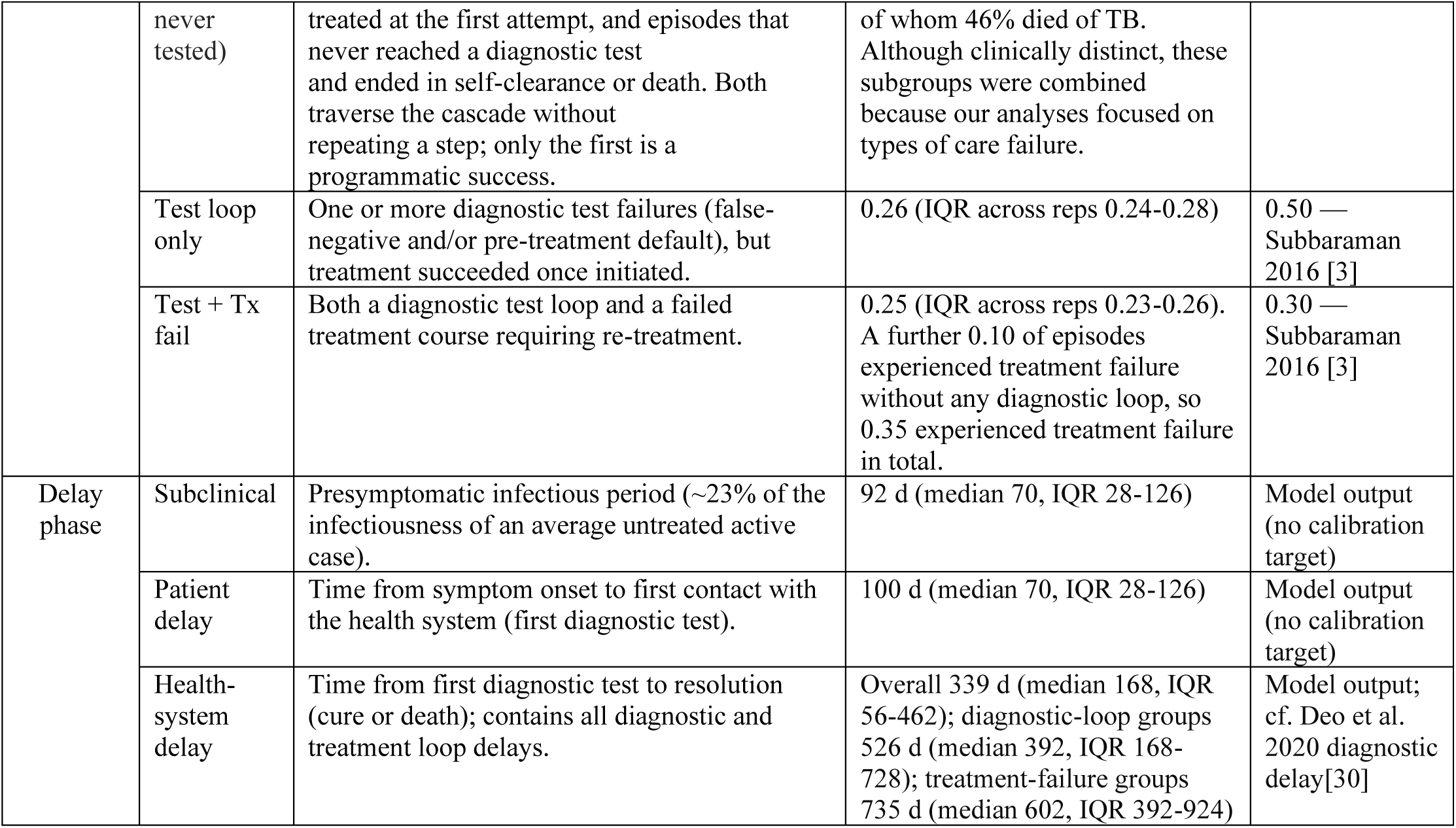
Definitions of loop-group classifications, delay phases, and resource-use metrics in the TB cascade model.

### Intervention scenarios

To evaluate the relative and combined effects of interventions on TB transmission dynamics, cumulative infectiousness, and healthcare resource utilization across care-seeking groups, we defined eight intervention scenarios, with all interventions implemented beginning in 2025 (Table 3). The baseline scenario represented the existing TB control strategy based on directly observed therapy short-course (DOTS) and current GeneXpert coverage. Three intervention components were evaluated individually and in combination: Active case finding (ACF), Improved diagnostics (Dx), and Treatment support (Tx), the last acting on both linkage to treatment and cure. The resulting eight scenarios were: Baseline, ACF, Dx, Tx, ACF+Dx, ACF+Tx, Dx+Tx, and ACF+Dx+Tx. Together, these scenarios enabled assessment of both incremental and synergistic effects of interventions targeting different stages of the TB care cascade.

**Table 3.**
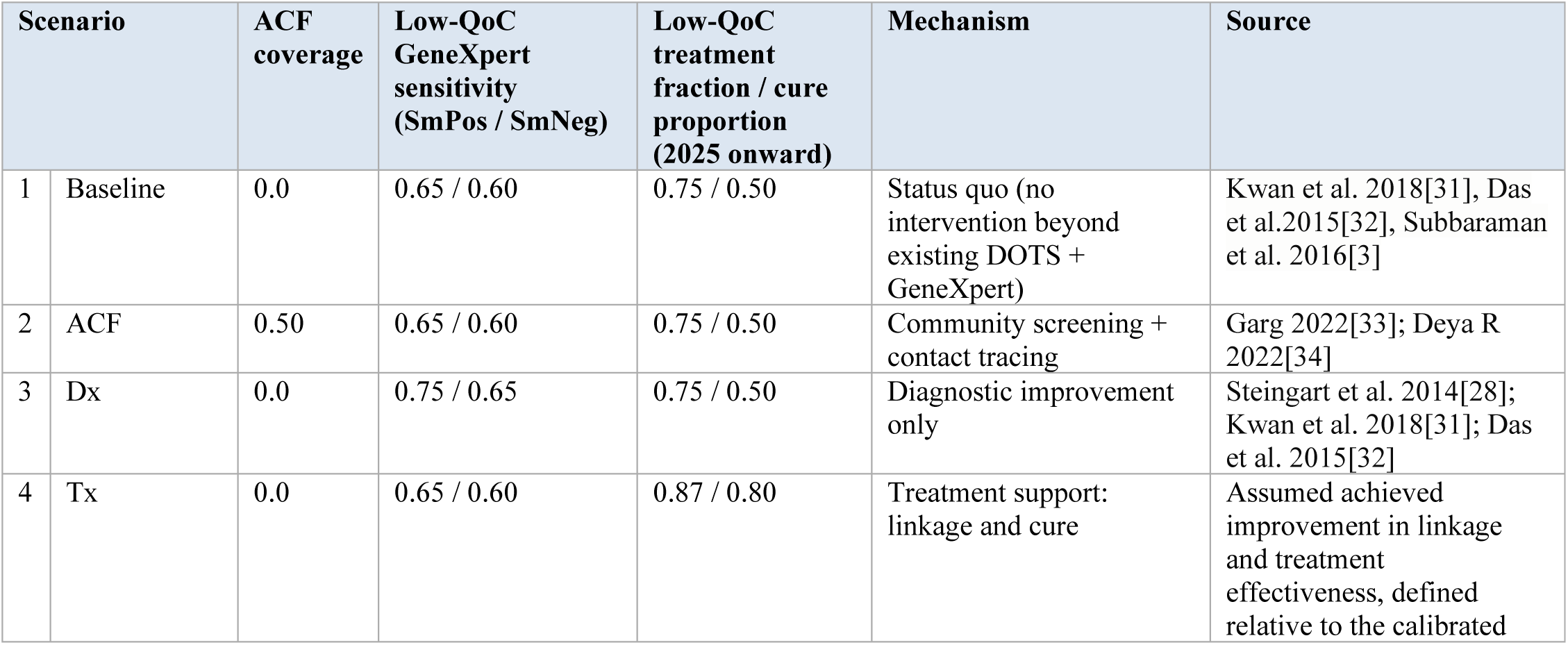
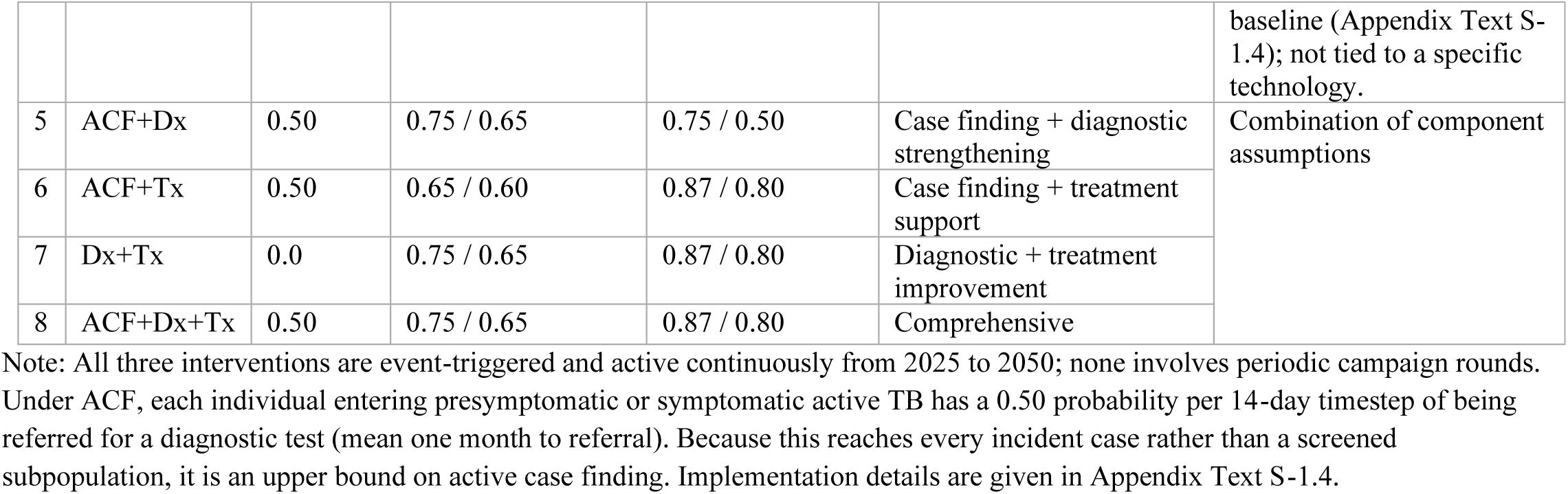
Intervention scenarios and corresponding EMOD parameter modifications.

### Outcome measurements

Primary outcomes were TB incidence, prevalence and mortality, cumulative infectiousness, active TB person-years, and lifetime diagnostic testing volume per TB-positive individual. Infectiousness attributable to subclinical disease, patient delay and health-system delay was translated into active TB person-years using the number of individuals in each care-seeking group, and the cumulative tests needed to reach a TB-positive diagnosis were capped at ten, the observed empirical ceiling. All outcomes were summarised across 100 stochastic replicates per scenario (98 completed; one in which TB died out before the intervention period was excluded, leaving 97 in every estimate). Intervention effects on incidence, prevalence and mortality were computed as the ratio of replicate means between scenario and baseline, with 95% confidence intervals from a replicate bootstrap, because TB deaths are a low-count channel on which per-replicate ratios are unstable; episode-level cascade quantities (Figure 4) were paired by random seed. Reductions were also tabulated across the three loop drivers, the infectiousness delay phases, and the three states composing health-system delay, separating a change in the number of loop events from a change in the time each event costs. Analyses used Python 3.10 with pandas 2.3, numpy 2.2, matplotlib 3.10 and scikit-learn 1.7.

### Calibration to epidemic burden and comparison with care-cascade benchmarks

We calibrated the model to six burden targets: WHO incidence estimates for 2000, 2010, 2015 and 2020, national survey prevalence for 2020, and WHO mortality for 2020[35]. Seven additional measures were reserved for plausibility checks: three loop-group fractions[3] and mean tests per case overall and within each group[36]. Published sequential cascade estimates were mapped to the model’s longitudinal groups for these comparisons (Appendix Text S-3.2). Incidence estimates for 2005 and 2023 were also withheld from calibration. Ten parameters were varied jointly, covering transmission, natural history, care-seeking timing and three historical trends (Appendix Table S-2). Lower-quality diagnostic sensitivity was fixed at its baseline value during calibration and served as the intervention lever in the diagnostic-improvement scenario. Treatment initiation followed a calibrated historical trend; the treatment-support scenario halved the remaining pre-treatment loss relative to its calibrated 2024 level.

Calibration used two-wave Bayesian history matching with Matérn-5/2 Gaussian-process emulators[37], requiring 4,500 simulations. The six target-specific emulators achieved cross-validated R² values of 0.84–0.95. An initial screening step excluded parameter combinations that failed epidemic viability checks, retaining 37% of the original parameter space. Applying the three-standard-deviation implausibility criterion to the six burden targets reduced the retained space to 5.76% of its original volume. The selected best-fitting parameter set matched all six burden targets within 0.3 standard deviations (maximum implausibility 0.27) and six of the seven cascade benchmarks within three standard deviations. The exception was the no-loop fraction: 0.39 in the model versus the published benchmark of 0.20, a difference of 3.2 standard deviations (Appendix Table S-5). Phase-specific delays provided additional plausibility checks but were not calibration targets because empirical estimates are affected by recall and survivor biases and often include only diagnosed patients. The full calibration protocol is provided in Appendix Text S-3 and Tables S-1 to S-6.

## Results

**Figure 2.**
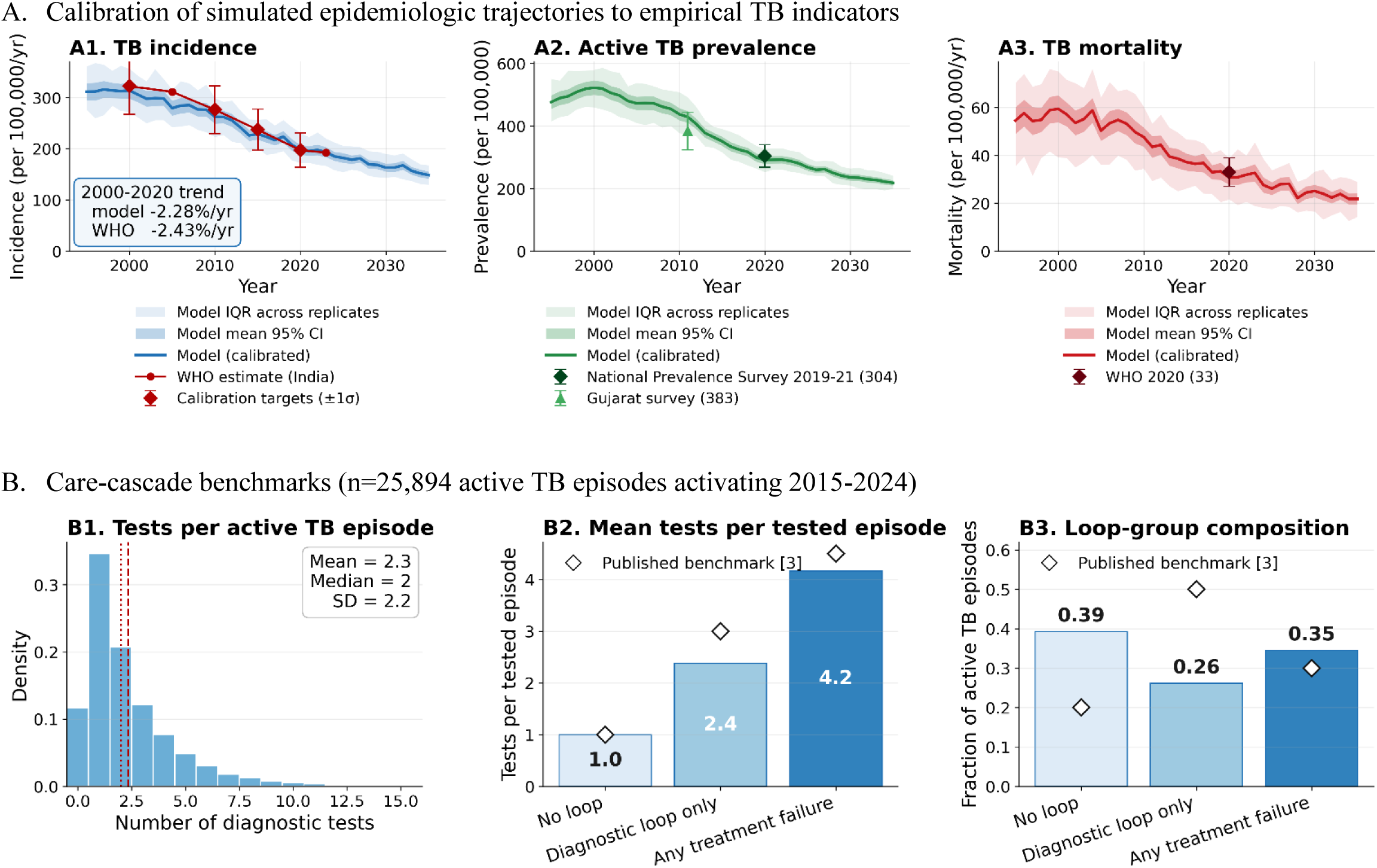
Model calibration and characterization of tuberculosis care-cascade burden Note: Baseline scenario of the production run (100 replicates) at the trend-calibrated parameter set. Incidence is calibrated to the WHO trajectory at 2000, 2010, 2015 and 2020 rather than to a single 2020 anchor; all six burden targets fall within 0.4 standard deviations of their anchors in the production run. Bands are the interquartile range across replicates and the 95% confidence interval of the mean. Interventions begin in 2025, so the baseline shown here is the no-further-improvement counterfactual against which the intervention scenarios are compared. In panel B3 the three groups are mutually exclusive: of the 0.35 of episodes with treatment failure, 0.25 also had a diagnostic loop and 0.10 did not. The share of episodes with at least one diagnostic loop is therefore 0.26 + 0.25 = 0.51, against the benchmark of 0.50.

The calibrated model reproduced India’s historical decline in incidence while matching prevalence and mortality benchmarks (Figure 2A). The annual incidence decline was 2.28%, compared with 2.43% in the WHO series, with good agreement in the held-out years. The model also captured the higher diagnostic testing intensity associated with more complex care pathways, although these measures were excluded from calibration (Figure 2B). Of the seven cascade benchmarks, the three loop-group shares and mean tests per tested episode overall and within each group (Appendix Table S-5), six fell within the prespecified tolerance of three standard deviations. Two gaps stand out. The no-loop share was 0.39 versus 0.20, the only benchmark outside tolerance (3.2 standard deviations). The diagnostic-loop-only share was 0.26 versus 0.50 (2.4 standard deviations); the benchmark counts every episode with a diagnostic loop, whatever followed. These two discrepancies reflect differences in definition rather than in the testing behaviour simulated. The no-loop group includes not only episodes treated at the first attempt but also episodes that were never tested or treated, which make up 30% of the group, so its share is larger than a cascade of treated patients implies. Conversely, a diagnostic loop followed by treatment failure is assigned to the treatment-failure group, which lowers the diagnostic-loop-only share: 51% of episodes had at least one diagnostic loop (0.26 + 0.25), in line with the benchmark of 0.50. Counting failures of either type, 61% of episodes (0.26 + 0.35) had at least one, against roughly 80% implied by the published cascade (0.50 + 0.30).

**Figure 3.**
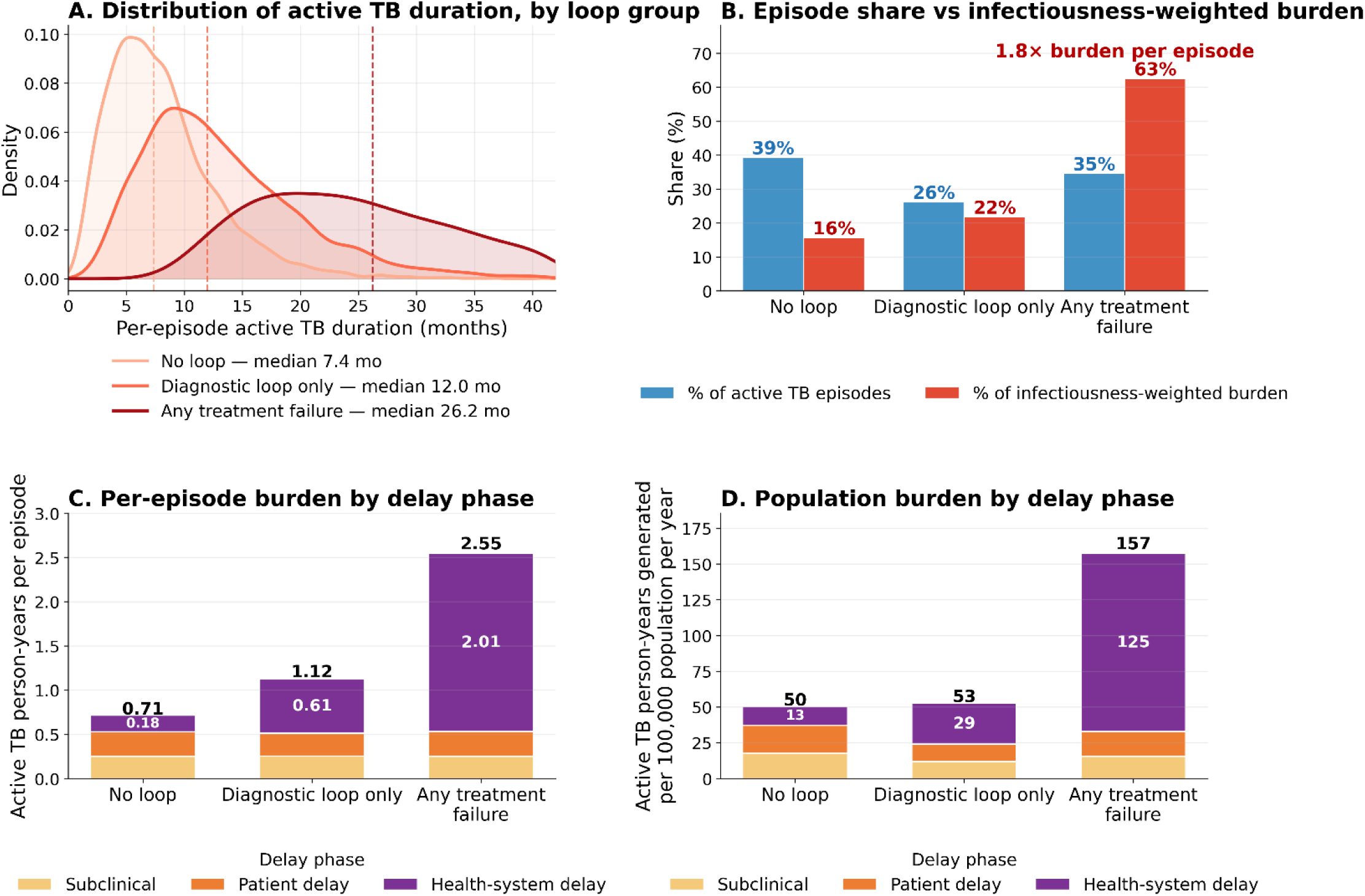
Contribution of loop groups to individual-level and population-level transmission burden Note: Baseline scenario, 100 replicates, 25,894 active TB episodes activating in 2015–2024 and followed to resolution. Panel A shows unweighted active TB duration; the median effective infectious time, weighted by infectiousness (presymptomatic phase × 0.23, symptomatic phase ×1, treatment phase × 0), is 3 months in the no-loop group, 8 months with diagnostic loops only and 16 months with treatment failure. In panels C and D the number inside each purple segment is the health-system-delay component, the person-time accrued after the first diagnostic test.

In the baseline scenario, decomposing the burden of episodes activating in 2015-2024 by care-loop group revealed that per-episode active TB duration increased markedly with cascade complexity (Figure 3A): episodes with no recorded loop event (NoLoop-only) remained in active disease for an unweighted median of 7.4 months, those with diagnostic loops only for 12.0 months, and those with treatment failure for 26.2 months — three and a half times longer. Weighted by infectiousness, which discounts the presymptomatic phase and excludes time on treatment, the corresponding median effective infectious times were 3, 8 and 16 months. This heterogeneity produced a pronounced Pareto-type disparity between episode share and infectiousness-weighted burden (Figure 3B): the treatment-failure group constituted 35% of active TB episodes yet generated 63% of the infectiousness-weighted burden — 1.8 times its episode share — whereas the NoLoop-only group represented 39% of episodes but contributed 16%. At the population scale (Figure 3D), the two looping groups generated 210 of the 260 active TB person-years per 100,000 population per year, four times the 50 generated by the no-loop group. Most of this burden arose after care entry: health-system delay, the interval from first diagnostic test to resolution, accounted for 64% of all active TB person-time and for four fifths of the treatment-failure group’s (purple segments in Figure 3D). The excess is therefore generated within the health system, in the part of the pathway that aggregate cascade indicators are least able to see.

**Figure 4.**
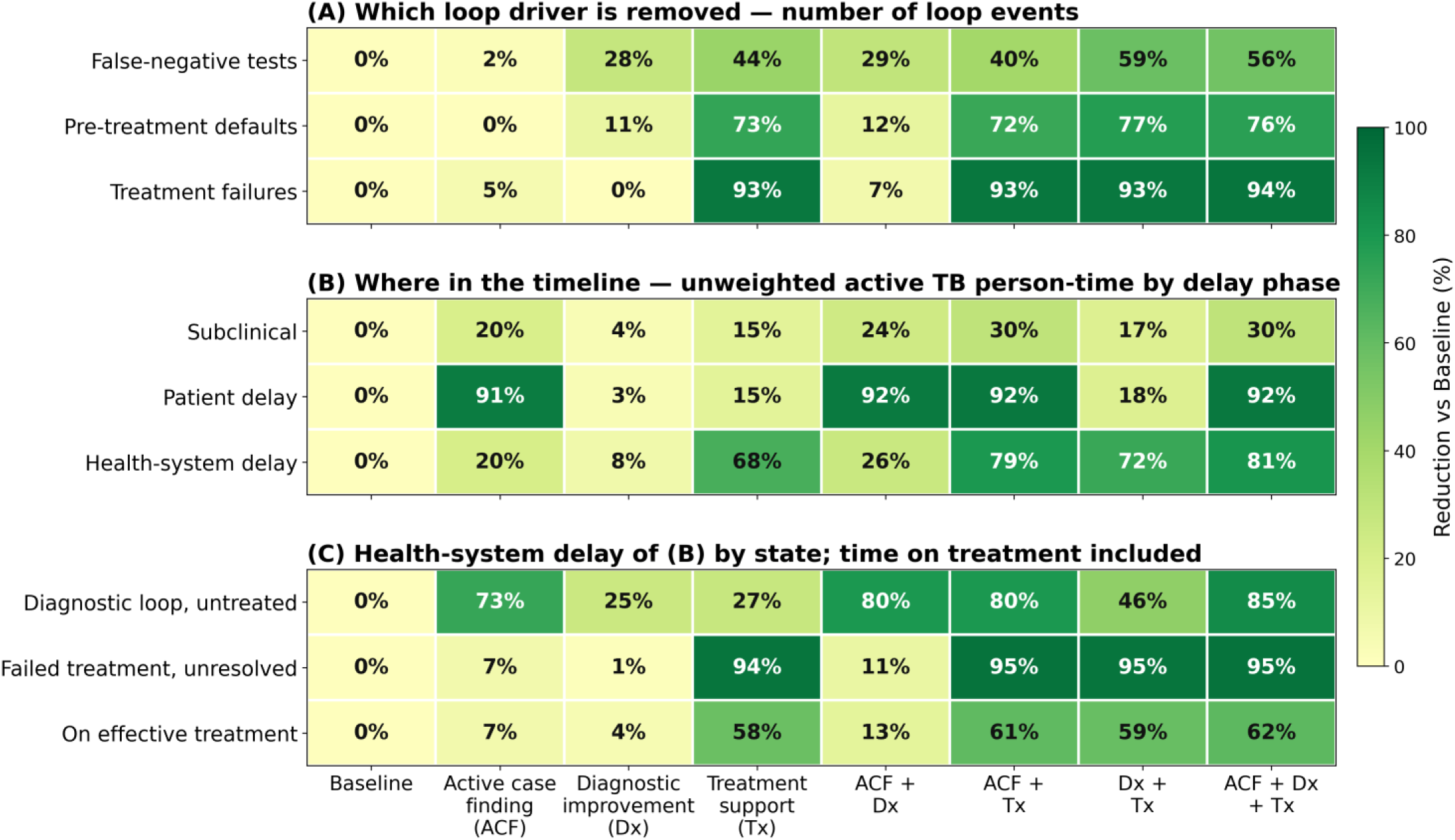
Three-lens view of intervention impact (% reduction vs Baseline). Note: Episodes activating from 2025 onward, 100 replicates, paired by random seed. Each cell is the percentage by which the scenario reduces that quantity relative to Baseline, as the mean of the paired per-replicate ratios. Panels B and C are unweighted active TB person-time, so time on effective treatment is included although it carries zero weight in the infectiousness-weighted burden of Figure 3. The largest paired 95% confidence interval half-width across all cells is 5.8 percentage points.

Figure 4 separates two ways an intervention can reduce active TB person-time: by preventing loop events and by shortening the time each event costs. The comparison uses unweighted person-time, including time on treatment. Active case finding worked almost entirely through time: it left loop-event counts nearly unchanged but reduced time spent in patient delay by 91% and in untreated diagnostic loops by 73%. Treatment support worked at the later stage, reducing health-system delay by 68%, primarily through a 94% reduction in time spent with unresolved treatment failure. Because the two act on different phases, combining them reduced person-time in health-system delay by 79%, approaching the 81% achieved by the full package. Diagnostic improvement alone had the smallest effect, reducing untreated diagnostic-loop time by 25% and overall health-system delay by 8%.

The comparison between active case finding and diagnostic improvement shows why event counts and person-time can diverge (Figure 4A and 4C). Diagnostic improvement reduced false-negative tests by 28% and active case finding by only 2%; yet active case finding reduced untreated diagnostic-loop time far more (73% versus 25%). The difference lay in the interval to repeat testing, which fell from 63 days at baseline to 17 days under active case finding but stayed at 61 days under diagnostic improvement. Active case finding thus shortened the untreated interval that follows a loop event, whereas diagnostic improvement reduced the number of events. Counting loop events alone would miss the larger benefit, which came from faster re-engagement: less time spent infectious between care encounters.

**Figure 5.**
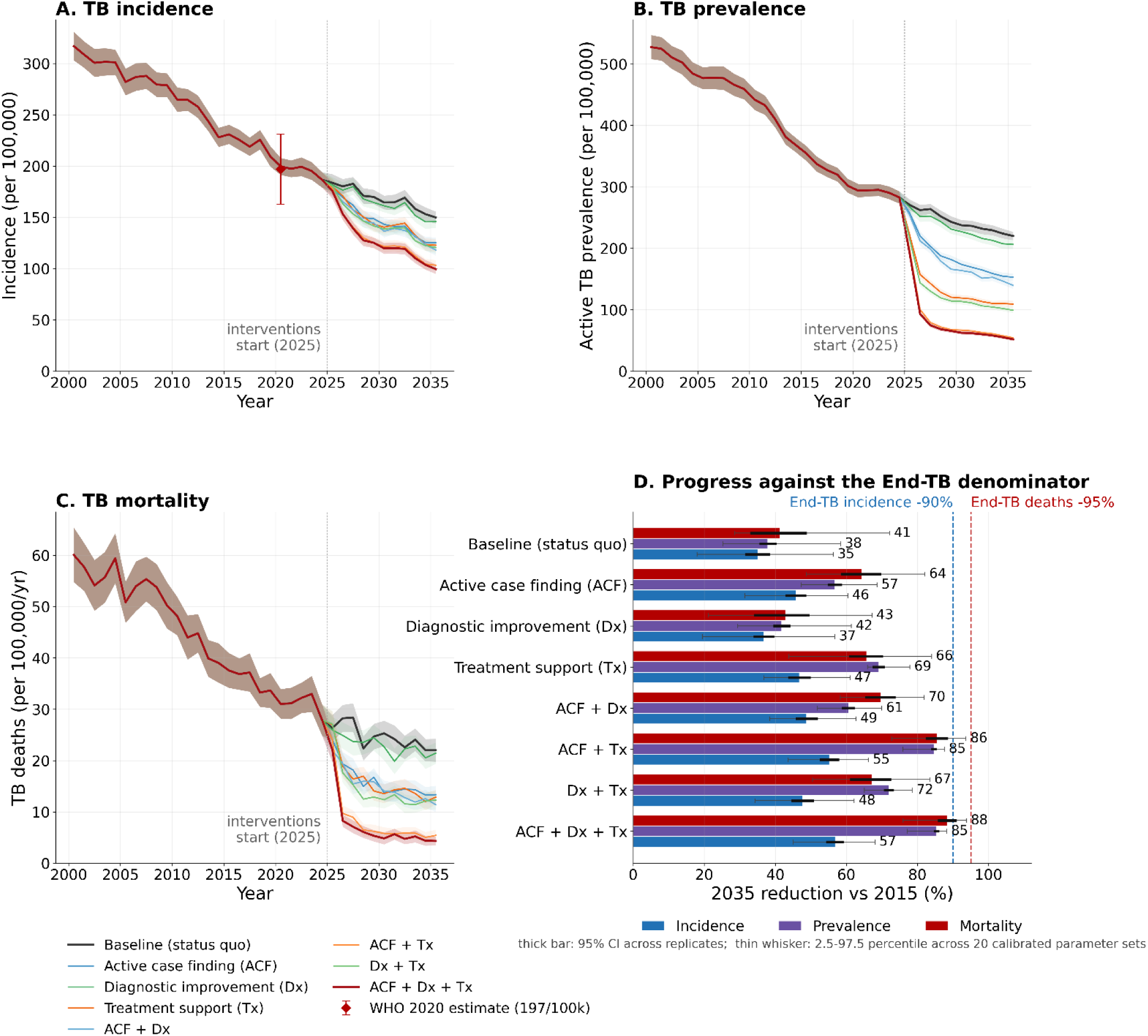
Projected progress toward the End-TB targets on TB incidence, prevalence and mortality Panels A-C show mean trajectories with 95% confidence intervals across stochastic replicates at the calibrated parameter set. Panel D gives the 2035 reduction against 2015 as the ratio of means, with the replicate bootstrap 95% CI (thick) and, as a thin whisker, the 2.5th-97.5th percentile range across 20 parameter sets drawn from the retained calibration region, each set measured against its own 2015 baseline (Appendix Text S-3.8); the ratio of means is used because TB deaths are a low-count channel on which the mean of per-replicate ratios is unstable. Sensitivity to further cascade-loop closure is shown in Appendix Figure S-2.

These changes in loop time translated into population effects (Figure 5). Incidence, prevalence and mortality all fell below the status-quo trajectory from 2025, with the largest separations under the combinations that included treatment support (Figure 5A–C). Measured from 2015, the baseline of the End TB targets (Figure 5D), the status quo alone reduced incidence by 35%, prevalence by 38% and deaths by 41%. Treatment support and active case finding reached similar reductions in incidence (47% and 46%) and mortality (66% and 64%) but differed on prevalence, where treatment support reached 69% against 57%, a difference largely preserved across the 20 parameter sets (thin whiskers). Diagnostic improvement alone moved none of the three measures appreciably beyond the status quo (37%, 42% and 43%). The full package reduced prevalence by 85% and deaths by 88%, close to the End TB mortality target of 95%, but incidence by only 57%, short of the 90% target in every parameter set; active case finding with treatment support came close to the same result (55%, 85% and 86%). This follows from where each acts (Figure 4): treatment support removes the years that patients with unresolved treatment failure remain sick, whereas earlier detection shortens a brief pre-diagnosis interval but, in doing so, also prevents the infections and deaths that would occur during it.

Further closure of the residual cascade losses showed where loop time still mattered (Appendix Figure S-2). Removing a further 25–75% of the residual losses raised the incidence reduction substantially wherever an intervention left loop time intact: from 2.7% to 20.7% under diagnostic improvement, from 16.5% to 32.6% under active case finding, and from zero to 23.9% with no other intervention, which on its own corresponds to a 50% reduction from 2015, the interim End TB milestone. The gains were smaller under treatment support (18.0% to 24.6%), which already removes most treatment-failure time, and negligible under the full package (33.6% to 34.6%), under which 85% of untreated diagnostic-loop time and 95% of unresolved treatment-failure time were already gone. Once the cascade was tightened to this point, the remaining gap to the 90% target no longer reflected care-cascade losses but infection acquired before the interventions, which generated about three quarters of projected cases in 2025–2035.

## Discussion

Our model of TB in India showed that the path through diagnosis and treatment matters for transmission, beyond whether a patient is eventually diagnosed or treated. Partitioning every active episode into four mutually exclusive loop groups showed that patients with the same WHO-defined treatment outcome can carry very different transmission burdens. Episodes with treatment failure accounted for 35% of all active TB episodes but 63% of the total infectiousness-weighted burden. By comparison, episodes with no recorded diagnostic or treatment failure accounted for 39% of episodes but only 16% of this burden. Median effective infectious time was approximately 16 months in the treatment-failure group, compared with 3 months in the no-loop group. The interventions divided along the same line: treatment support and active case finding reduced 2035 incidence comparably (18% and 17%), improved test sensitivity alone did not (3%), and the full package reached 57% from 2015, short of the 90% End TB target. This suggests two practical consequences. First, the treatment-failure group marks the care processes where improvement is most needed: shortening those trajectories would reduce transmission and at the same time relieve the repeat testing and repeat treatment that the same patients consume. Second, because many patients in this group are eventually cured and recorded as treatment successes, endpoint-based indicators do not distinguish them from patients treated at the first attempt; aggregate treatment-success rates should therefore be interpreted alongside measures that capture the trajectory[38]. Grouping episodes by loop history provides such a measure: it identifies which subgroup carries the transmission, which interventions act on which subgroup, and where in the cascade the remaining time can be removed.

The main insight is that counting care events alone does not reveal how much infectious time they create or prevent. The consequences of a false-negative test depend on both its frequency and the untreated interval before the next encounter. Patients with the same number of failures can therefore accumulate very different infectious burdens. In our simulations, active case finding reduced false-negative tests by only 2% but shortened repeat-testing intervals from 63 to 17 days, reducing untreated diagnostic-loop time by 73%. Diagnostic improvement prevented more false-negative tests (28%) but left the interval largely unchanged at 61 days, reducing diagnostic-loop time by 25%. Preventing failures and shortening the delays that follow them are thus distinct ways to reduce infectious burden. The smaller diagnostic effect also reflects the scenario’s limited scope: sensitivity improved without changes in referral, treatment linkage, or repeat-testing intervals[39]. Broader diagnostic strengthening could address these processes alongside test accuracy[40–44]. Similarly, the benefits of active case finding depend on the disease trajectories of those detected, including possible progression or regression of subclinical disease[45], and on the care they subsequently receive. Earlier detection can shorten infectious time, while timely linkage and effective treatment help sustain that benefit[30, 46]. Whether a particular tool delivers such improvements is an implementation question: digital adherence technologies did not improve treatment outcomes in recent pragmatic trials[47]. Active case finding and treatment support therefore acted through complementary mechanisms: the former shortened delays before and between diagnostic encounters, while the latter reduced time with unresolved treatment failure. Their combined effect illustrates why intervention evaluation should measure both the frequency and duration of cascade failures.

Malhotra and colleagues[48] evaluated active case finding against the course each detected person would have followed under routine care, in which some would have recovered spontaneously and others been diagnosed later, so that the benefit of detection is the infectious time it removes relative to that counterfactual. Trial evidence fits this framing: in a systematic review of community-based case finding[49], prevalence fell where sputum testing was offered to whole communities in repeated rounds, as in Vietnam, but not where a single mobilisation-based round reached fewer people, as in South Africa and Zambia; the difference lies in how much infectious time each approach actually removed. Kendall and Dowdy[17] applied the same reasoning at the individual case, separating the benefit to the person detected from the benefit to others. The first depends on timing: detection must come before routine diagnosis would have occurred and before death or sequelae become unavoidable, a window that is often narrow. The second accrues throughout the interval that detection removes, and in their analysis accounted for 65% of the DALYs averted by mass screening. The value of finding a case therefore lies less in the diagnosis itself than in who is found and how much infectious time is thereby removed. These studies measure the infectious time removed before a first diagnosis. We applied the same accounting to the time that accrues after care entry, through false-negative tests, pre-treatment loss and treatment failure, which in our model held 64% of all active TB person-time. Heterogeneity in who is detected has so far been described biologically, by smear or symptom status; loop histories add the heterogeneity of care pathways, and together the two explain differences in cumulative infectious burden. Loop histories separate prompt treatment success from eventual success after prolonged interruptions.[38] [50] Linking notification, repeat-testing, and treatment-outcome records could make these pathways visible in routine data, records across providers[51][52]; such linkage would show programmes where delays and repeated failures occur and which care processes require improvement[53].

The remaining gap to the End TB incidence target partly reflects when infections occurred. Multi-model analyses suggest that existing interventions alone may be insufficient to achieve global TB targets[54]. Our projections help explain one source of the remaining gap to the End TB incidence target: approximately three quarters of cases during 2025–35 arose from infections acquired before intervention implementation. The full package reduced new infections by 82% and rapid progression by 78%, but reactivation by only 5%; reactivation accounted for 96% of residual incidence in 2035. By then, prevalence under the full package was 85% lower than in 2015, compared with a 57% reduction in incidence. This contrast is consistent with care improvements shortening disease duration while having less effect on disease arising from older infections. Preventive therapy or suitable vaccines could complement care improvements by preventing progression among already-infected populations. Presymptomatic disease accounted for half of residual active TB prevalence, highlighting the potential value of detection and treatment before symptom-driven care.

The different responses of incidence, prevalence, and mortality reflect how cascade strengthening affects disease duration, survival, and transmission over time. Earlier detection and effective treatment can shorten active disease and reduce mortality, whereas incidence benefits from prevented transmission emerge more gradually and are moderated by disease arising from earlier infections. The prevalence-to-incidence ratio can help summarise these dynamics, although it approximates mean disease duration only under steady-state conditions and with comparable disease definitions. In our projections, status-quo incidence declined by 35% between 2015 and 2035, while this ratio remained nearly unchanged, from 1.54 to 1.47 years. By contrast, the full intervention package reduced the ratio to 0.52 years in 2035, as prevalence fell much more sharply than incidence. This pattern is consistent with cascade strengthening shortening the time people lived with active TB, beyond the reductions accompanying the baseline epidemic decline. The faster prevalence response—within two years in these projections—also highlights the importance of the evaluation timeframe: incidence alone may understate the near-term benefits of cascade investment within a five-year planning cycle. Assessing prevalence and mortality alongside incidence can therefore provide a fuller account of intervention impact. Because prevalence surveys are infrequent and costly, linked patient records could provide complementary measures of care-related duration, including repeat-testing intervals, time to effective treatment, and re-engagement after failure.

For programme monitoring, the practical implication is to add measures of time and continuity to existing counts[38]. Useful measures include time from first contact to effective treatment, intervals between repeat tests, and time from treatment failure to re-engagement. Separating delays before and after care entry would help distinguish access barriers from problems within the health system, although subclinical and undiagnosed disease would still require additional data sources. These measures would also support empirical validation of the care trajectories represented in the model. Establishing whether the model reproduces observed delays, interruptions, and re-engagement patterns across settings is essential to assessing the broader applicability of the care-cascade framework and the transferability of effect sizes estimated under this India-specific parameterization. Prevalence surveys could provide complementary evidence. Under a strengthened care cascade, half of the remaining prevalent disease was presymptomatic. Symptom-based detection alone may therefore miss a growing share of disease as programmes improve. Across our 2035 scenarios, 18–46% of prevalent disease occurred among people receiving treatment. Reporting prevalence separately for people on treatment and people not yet treated would show how much prevalent disease is untreated, and therefore still infectious, and how much is disease that persists during treatment. It would also make prevalence-to-incidence ratios easier to compare across settings, with the caveat that such ratios approximate disease duration only when incidence is stable and prevalence and incidence use the same case definition.

Several limitations should be considered when interpreting these findings. First, assumptions about TB natural history affect estimates of future disease arising from earlier infection[56, 57]. The model allows spontaneous resolution of untreated active TB but assumes that latent infection persists with a constant hazard of slow progression. Untreated disease duration and case fatality were also fixed rather than calibrated. These assumptions influence how long new cases continue to arise from pre-existing infection after interventions are introduced. Thus, although interventions can rapidly reduce the number of people with active TB, continued progression from earlier infection may cause incidence to decline more slowly than prevalence in our projections. The magnitude and duration of this difference depend on the natural-history assumptions. Incorporating evidence of infection clearance and disease regression[56, 57] into recalibrated models could refine estimates of the population at risk and the timing of disease onset, helping identify whom complementary interventions should reach. Second, simplified assumptions about care quality, repeat testing and delays in care seeking and re-engagement may affect the relative benefits of preventing failures versus shortening delays. Diagnostic or treatment-failure groups comprise 61% of modelled episodes versus roughly 80% in the published cascade, although definitions and denominators differ. If preventable failures are underrepresented, the benefits of addressing them may be underestimated. Conversely, the scenarios assume that improvements are achieved from 2025, with case finding reaching all incident cases, including presymptomatic disease. Early reductions in prevalence and mortality may therefore exceed those achievable through gradual programme expansion. The net effect of these opposing influences on ACF estimates remains uncertain; realised benefits will depend on coverage, continuity of care and implementation capacity.

Third, calibration constrained parameter combinations more tightly than individual values. Only base infectivity and the slow-progression rate were narrowed appreciably; the other eight free parameters retained roughly 90% of their prior range (Appendix Table S-7). This is expected when six aggregate burden targets are asked to constrain ten parameters that can compensate for one another, for example a higher base infectivity offset by a lower slow-progression rate, and history matching discards parameter sets inconsistent with the data without ranking those that remain. The care-cascade benchmarks could not break these trade-offs: cascade composition is set mainly by inputs held fixed, such as test sensitivity, pre-treatment loss and treatment failure, and shifted so little across parameter sets consistent with the burden targets, less than half the benchmark tolerance, that the benchmarks could not discriminate among them. The calibrated values should therefore be read as one plausible combination rather than as estimates of individual parameters. A wide retained range for the operational parameters is also realistic, since care-seeking delays and long-term trends vary across states, providers and periods. The analysis also does not capture uncertainty in fixed parameters or values outside the prior bounds, including a presymptomatic phase longer than the assumed six months. Although these uncertainties affect absolute burden estimates, intervention effects were consistent across the twenty retained parameter sets, which supports the main conclusions within the tested space, including the limited effect of diagnostic improvement alone on incidence.

Future studies may select parameters for calibration according to the strength of external evidence, their influence on relevant outcomes and whether the available data can distinguish their effects[58][59]. This study gave priority to burden targets, because care-cascade indicators vary widely between settings, and fixed the inputs that determine cascade composition, such as test sensitivity, pre-treatment loss and treatment failure. Where setting-specific data on these processes can be collected, such as linked records of care-seeking, testing and treatment, and survey evidence on subclinical disease, those inputs could be calibrated, and burden and cascade targets weighted according to the research question. Such data could separate processes that produce similar aggregate burden[59] and distinguish variation across settings from uncertainty in average values; where they are unavailable, external evidence can enter as informative priors [60]. When estimates of individual parameters are required, history matching could be followed by a likelihood-based step, such as approximate Bayesian computation or an emulator-based posterior, that accounts for observation and simulation uncertainty [61][62]. Greater precision would depend on more informative data and an appropriate model structure rather than on additional calibration runs[63, 64].

## Conclusion

This study demonstrates the value of evaluating TB care through both the frequency of diagnostic and treatment failures and the infectious time they create. In our model, these measures clarified why interventions at different stages of care produced distinct effects on incidence, prevalence, and mortality. Monitoring repeat testing, treatment failure and the time to starting or resuming effective treatment would complement conventional cascade indicators and help identify where care improvements are most needed. Linking encounters across providers through case-based platforms could support this approach. Evaluating these care trajectories alongside population outcomes would provide a stronger basis for assessing earlier detection, diagnostic improvement, and treatment support, and for informing programme priorities and investment.

## Data availability

All model data and source code used in this study are publicly available at: https://github.com/UConn-Health-Disease-Modeling/tb-india-loop-cascade-abm. Raw simulation outputs (8–45 GB per experiment) are not distributed because of their size; they can be regenerated with the notebooks provided. The model runs on EMOD-TBHIV (DTK v2.21), open-source software developed by the Institute for Disease Modeling (https://github.com/InstituteforDiseaseModeling/EMOD), through the emodpy-tbhiv Python package (version 1.3.1, https://github.com/InstituteforDiseaseModeling/emodpy-tbhiv), which downloads the model executable; documentation is available at https://docs.idmod.org. Package versions are listed in the repository.

## Ethics statement

This study used a simulation model informed by published, aggregate data from the WHO Global TB Programme, the National TB Prevalence Survey of India 2019–21 and the peer-reviewed literature. No individual-level or identifiable data were used, and ethics approval was therefore not required.

## Author contributions

SC and YJ conceived the study and built and calibrated the model. SC advised on study design and the care-cascade framework and supervised the work. YJ ran the calibration and scenario simulations, performed the analyses, prepared the figures and wrote the first draft. JB developed the EMOD-TBHIV platform components used here, advised on model configuration and supported model calibration. SS and EW provided critical feedback on model validation and data interpretation, and EW provided the simulation platform and computing resources. KB supported the care-cascade group analysis and the simulation runs. All authors reviewed and edited the manuscript and approved the final version.

