## Supplementary Document for "Care-cascade failures as a determinant of infectious burden in tuberculosis: an agent-based modelling study in India"

Affiliations:
1. Department of Public Health Sciences, School of Medicine, University of Connecticut, Farmington, CT, USA
2. Institute for Disease Modeling, Gates Foundation, Seattle, WA, USA
3. Independent researcher, San Francisco, CA, USA
4. Research Computing, Information Services & Technology, Boston University, Boston, MA, USA

### Text S-1. Model methods

#### S-1.1 Population parameterization and simulation

The EMOD-TBHIV simulation population was initialized from a single modelled node with India-specific demography. Age- and year-specific all-cause mortality was applied from an India-specific series spanning 1991–2018[1] and the crude birth rate was scaled so that the simulated population reproduces India's growth and ageing over the simulation period. Heterogeneity in transmission and in care was represented through individual properties rather than through geographic nodes, so the model contains no explicit rural–urban structure or migration. Age-specific contact patterns were parameterized using the India Prem contact matrix, aggregated into four age groups: 0–19, 20–39, 40–59, and ≥60 years[2]. To approximate heterogeneity in TB burden without explicit geography, individuals were assigned to low-, medium-, or high-prevalence strata in approximately equal thirds, informed by a classification of 28 Indian states, with strongly assortative contact between strata (within-stratum weights 2.9–3.5 against 0.1 between strata). Quality of care was assigned as a separate and independent individual property (higher- vs lower-quality stream; Table 1) that governs each individual's cascade parameters without altering contact patterns.

The statistical simulation population increased with India's demographic trajectory from approximately 20,200 at model initiation in 1850 to 178,600 in 1990, 297,400 in 2020, and 502,400 by 2049. The 2020 simulation population represented India's approximately 1.38 billion population at a scaling ratio of approximately 1:4,600. With a 5% individual sampling rate, each computational agent corresponded to approximately 93,000 individuals. The simulation ran from 1850 through 2050, providing 147 years of burn-in before introduction of the DOTS programme in 1997; calibration outputs are presented primarily for 2000–2035. Across the production replicates, the model generated approximately 334,600 active TB activation events over the full simulation period, including approximately 2,874 events within the 2020 ±6-month calibration window.

#### S-1.2 TB natural-history parameterization

Following infection, individuals entered either a fast- or slow-progressing latent state. Children aged <15 years had a 3% probability of fast progression to active TB, whereas adults aged ≥15 years had a calibrated probability of 7.6%.[3] Fast progressors left the latent state after a mean of approximately 2 months. The remaining individuals entered the slow-progressing latent state, with a calibrated annual reactivation rate of 0.30%, corresponding to an approximately 14% cumulative probability of activation over 50 years. Latent infection was assumed to be non-infectious, was not associated with additional mortality, and did not self-clear. Latent individuals nevertheless remained exposed to the force of infection, at approximately 32% of the susceptibility of never-infected individuals (Table S-2). Exogenous reinfection was enabled, so an infection acquired during latency re-entered the progression draw rather than being absorbed into the existing latent state: a reinfected slow progressor could therefore progress on the fast pathway rather than on the 0.30% annual reactivation hazard. Individuals carried only one infection at a time, so reinfection replaced rather than accumulated. This is the route by which transmission intensity feeds back into progression.

Progression to active TB occurred through a pre-symptomatic phase with reduced infectiousness before symptomatic disease. The pre-symptomatic phase had a calibrated mean duration of approximately 85 days, so fast progressors reached symptomatic disease after approximately 4.7 months on average. Neither additional mortality nor spontaneous regression was assumed during the pre-symptomatic phase. Symptomatic active TB was classified as smear-positive pulmonary, smear-negative pulmonary, or extrapulmonary disease (65%, 25%, and 10% of adult cases, respectively). Smear-positive pulmonary TB had the highest infectiousness; smear-negative pulmonary TB was assumed to be approximately 35% as infectious as smear-positive disease, the pre-symptomatic phase 16.6% as infectious, and extrapulmonary TB non-infectious.

In the absence of treatment, smear-positive disease resolved spontaneously or ended in TB-related death after a mean of approximately 2.75 years, with a case-fatality ratio of 0.65. The relative probabilities of spontaneous resolution and TB-related mortality varied by clinical presentation but not by age, mortality being scaled by 0.30 for smear-negative and 0.15 for extrapulmonary disease; across the adult smear distribution this corresponds to a mean untreated duration of approximately 3.6 years and an overall case-fatality ratio of approximately 0.53. Following successful treatment, individuals transitioned to a recovered state. Previously infected individuals, whether latent or recovered, had approximately 68% reduced susceptibility to reinfection compared with never-infected individuals. Individuals who subsequently relapsed were tracked separately from those achieving durable cure and re-entered the active-TB and care-seeking pathway. Natural-history parameters were held constant throughout the simulation period.

Figure S-1. Natural history of tuberculosis and individual care-cascade trajectories in the model

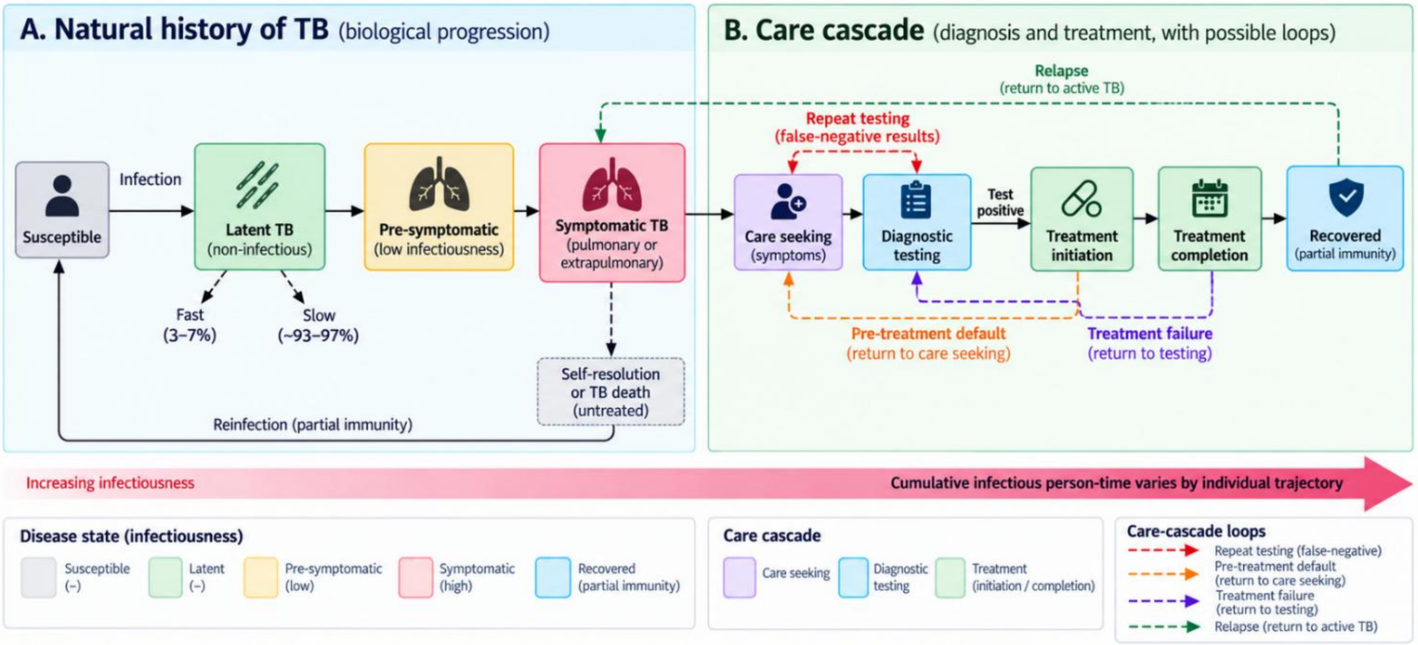

Note: individuals progress from infection to disease and may experience repeated diagnostic and treatment episodes, producing heterogeneous infectious durations. Panel A shows the biological progression and Panel B the care cascade, with the four loop pathways that return a patient to an earlier stage: repeat testing after a false-negative result, pre-treatment default, treatment failure, and relapse.

#### S-1.3 Care access and historical intervention parameterization

Symptomatic individuals were assigned to higher- or lower-quality care pathways representing heterogeneity in access to and quality of TB services in India (15% and 85% of the population, respectively; Table 1). The two pathways differed in diagnostic sensitivity and in treatment efficacy, but not in the time to care seeking. The higher-quality pathway had a smear-positive diagnostic sensitivity of 0.70 before GeneXpert and 0.75 after, and a treatment cure proportion of 0.80; the lower-quality pathway had sensitivities of 0.60 and 0.65 and a cure proportion of 0.50. Time to care seeking was instead represented by two subgroups common to both pathways, with calibrated mean delays of approximately 14 and 158 days. Parameter values for care-seeking delays, diagnostic performance, and treatment outcomes were derived from published India-specific literature (Table S-9).

The simulation incorporated historical changes in TB diagnosis and treatment. Following a pre-DOTS period, DOTS was introduced in 1997, with the treatment-initiation fraction starting at 0.70 and rising by a calibrated 0.17 percentage points per year to 0.75 by 2024, applied equally to both care pathways. GeneXpert availability was introduced from 2012 and scaled up over the following years. From 1997 to 2024, the per-step probability of care seeking improved by a calibrated 1.65% per year, a cumulative factor of 1.56 by 2024, and a secular non-cascade decline in transmission was applied as a node infectivity multiplier falling by a calibrated 1.28% per year to 0.71 by 2024. Upon care seeking, symptomatic individuals underwent sputum smear microscopy or GeneXpert testing according to test availability; diagnostic outcomes were determined by test-specific sensitivity, and individuals with a positive result proceeded to treatment. All three improvement trends were held at their 2024 values thereafter, so that historical service changes through 2024 define the calibrated baseline against which all interventions introduced from 2025 onward were evaluated.

##### S-1.4 Intervention scenarios from 2025

All three interventions are implemented as event-triggered interventions that remain active from the intervention start (2025) to the end of the simulation (2050); none is a periodic campaign. Active case finding is a health-seeking listener triggered by entry into presymptomatic or symptomatic active TB: each individual is referred for a diagnostic test with probability 0.50 per 14-day timestep, a mean of one month from activation to referral, with 75% of incident cases referred within one month and 94% within two. Referred individuals enter the existing diagnostic streams and remain subject to their sensitivities, so false-negative results and pre-treatment defaults are not removed; this is why active case finding increases the number of loop events while shortening the interval each costs (main text, Figure 4). The optimism of the scenario lies in its reach rather than its speed: it applies to every incident case in the population, including presymptomatic disease, whereas a real programme screens a subpopulation at intervals. Diagnostic improvement raises the lower-quality stream's test sensitivity to the higher-quality value from 2025, leaving the higher-quality stream unchanged. Treatment support acts on both halves of treatment in the lower-quality stream: it halves the residual pre-treatment loss relative to the calibrated 2024 initiation fraction (0.746 → 0.87), and raises the cure proportion to the higher-quality value (0.50 → 0.80), implemented as a separate regimen distributed from 2025 so that earlier years are unaffected (Table S-9).

#### Text S-2. Sensitivity of intervention impact to further cascade-loop closure

We tested whether further reductions in cascade failures could substantially increase the impact of the full intervention package. Reducing the residual loss at each loop driver by a further 25%, 50%, or 75% yielded 2035 incidence reductions of 32%, 35%, and 35% relative to the 2035 baseline, compared with 34% for the original package. These differences were small relative to replicate variation. The package already removed 85% of untreated diagnostic-loop time and 95% of unresolved treatment-failure time (main text, Figure 4C), leaving limited scope for further gains from these pathways. Larger gains occurred in scenarios that initially left more loop time unresolved: at the greatest additional closure, loop closure alone reduced incidence by 24%, and diagnostic improvement with additional closure achieved 21%, compared with 3% for diagnostic improvement alone. Within the tested scenarios, further closure therefore did not remove the remaining incidence gap. This is consistent with the main-text finding that approximately three quarters of TB episodes arising in 2025–2035 originated from infections acquired before the interventions.

Figure S-2. Sensitivity of intervention impact to further cascade-loop closure

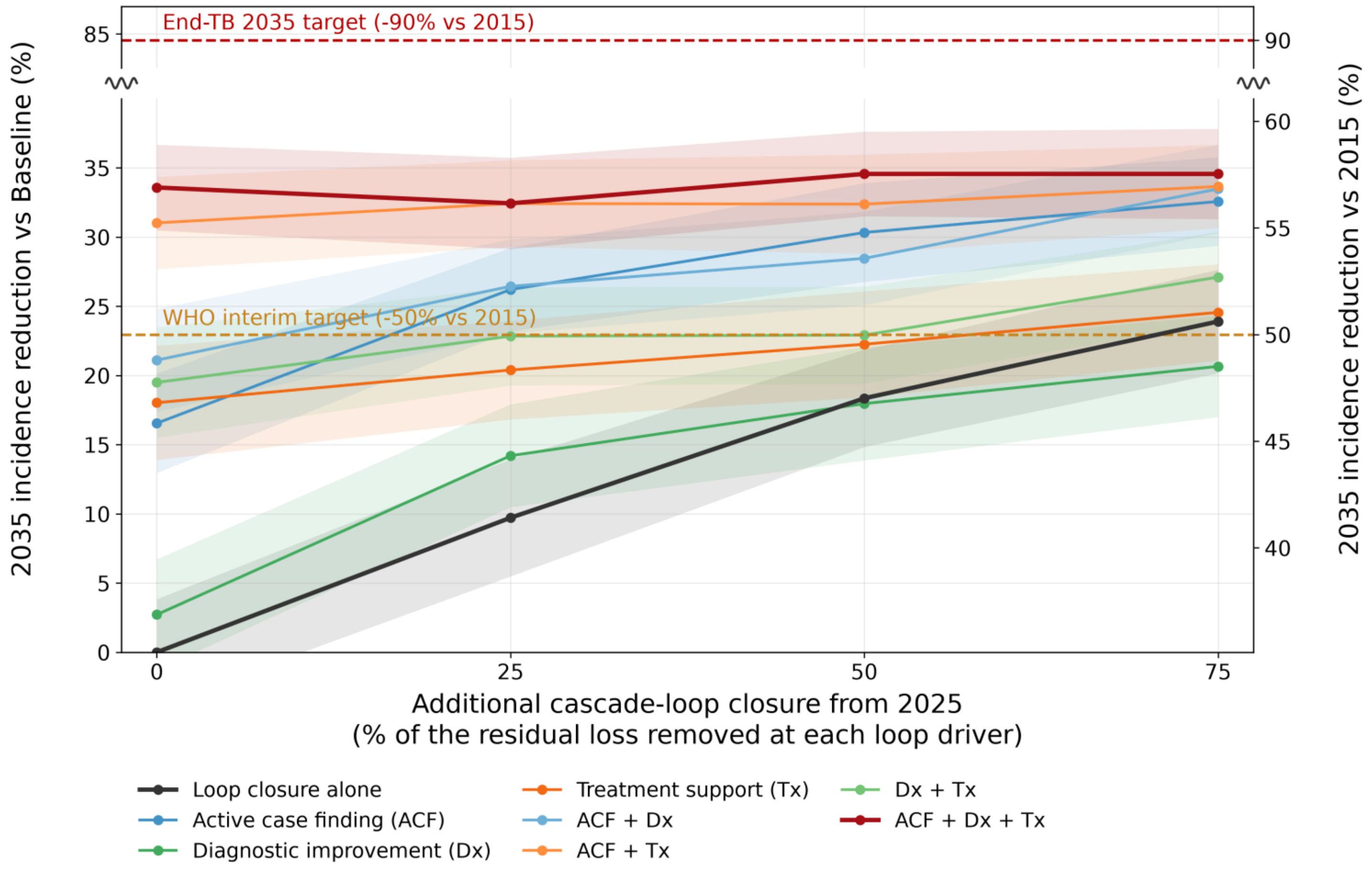

Note: From 2025, an additional 25%, 50%, or 75% of the residual loss at each loop driver is removed (24 design points × 100 replicates); zero closure is the main projection. Shading shows replicate-based 95% confidence intervals. The left axis gives incidence reductions relative to the 2035 baseline; the right axis gives reductions relative to 2015. Because baseline incidence falls to 65% of its 2015 level, 50% and 90% reductions from 2015 correspond to approximately 23% and 85% on the left axis. The axis break separates the projected effects from the 90% target.

#### Text S-3. Calibration via Bayesian history matching

This section describes the calibration of the EMOD-TBHIV agent-based model to thirteen empirical targets observed in the Indian TB epidemic (Table S-1): six burden targets used as emulator-gated calibration targets - four points on the WHO incidence series for India[4] plus 2020 prevalence and mortality - and seven care-cascade targets (three composition fractions and four mean-tests-per-case statistics) reserved as benchmarks for assessing care pathways not directly targeted by the fit. We use Bayesian history matching with Gaussian-process (GP) emulators (Andrianakis et al. 2015)[5] and the multi-wave protocol applied to TB-cascade calibration by Vesga et al. (2019)[6] and McCreesh & White (2018)[7]. History matching iteratively rules out regions of parameter space inconsistent with the observed data, without estimating a posterior probability distribution for the parameters, and treats structural model-data discrepancy as a distinct source of uncertainty alongside observational noise and emulator approximation error.

#### S-3.1 History matching framework

For each of the six burden calibration targets $i \in \{1, \dots, 6\}$ and parameter point x in the prior space, implausibility is

$$I_i(x) = \frac{|\mathbb{E}[f_i(x)] - z_i|}{\sqrt{\sigma^2_{\text{total},i} + \text{Var}_{\text{emul}}[f_i(x)]}}, \; x \in \text{NROY} \Leftrightarrow \max_{i \in \{1,\dots,6\}} I_i(x) \leq 3$$

Here, f denotes a model output, E its emulator-predicted mean, and Var_emul the prediction variance; z denotes the empirical target. The combined standard deviation σ_total includes observational and model-discrepancy uncertainty (Section S-3.3). A parameter combination is retained in the not-ruled-out-yet (NROY) region only if its implausibility is no greater than 3 for every burden target. Thus, it must be compatible with all six targets simultaneously. The seven cascade benchmarks do not determine which combinations are retained (Section S-3.7).

A combination is ruled out if it fails even one burden target. Successive waves sample the retained region and refit the emulators to improve screening. NROY identifies combinations that have not been ruled out under the specified uncertainty allowances; it does not establish that these combinations are uniquely correct.

#### S-3.2 Calibration targets

Thirteen empirical targets are listed in Table S-1. Incidence is matched along a trajectory rather than at a single year: the WHO Global TB Programme series for India (World Bank mirror of indicator SH.TBS.INCD)[4] supplies anchors at 2000, 2010, 2015 and 2020, and 2005 and 2023 are held out of the fit as a check on unused years. The 2020 prevalence anchor comes from the Indian National TB Prevalence Survey 2019-2021 and the 2020 mortality anchor from the WHO Global TB Report. Seven care-cascade targets - three composition fractions and four tests-per-case statistics - are taken from Subbaraman et al. (2016)[8] and Cazabon et al. (2017)[9].

The burden targets cover both the level and the historical trajectory of the epidemic. Matching several incidence years helps exclude parameter combinations that reproduce the 2020 level but imply a flat or rising epidemic over the preceding two decades. The care-cascade benchmarks provide a separate assessment of whether the fitted model also reproduces the observed structure of care pathways.

Table S-1. Empirical anchors and uncertainty allowances for calibration and benchmarking

| | Target | Family | Role | Anchor z | $\sigma_{obs}$ | $\sigma_{disc}$ | $\sigma_{total}$ | Source |
|---|---|---|---|---|---|---|---|---|
| 1 | TB incidence 2000 | Burden | Calibration | 322 | 43 | 34 | 54.7 | WHO Global TB Programme |
| 2 | TB incidence 2010 | Burden | Calibration | 276 | 37 | 29 | 46.9 | as above |
| 3 | TB incidence 2015 | Burden | Calibration | 237 | 32 | 25 | 40.3 | as above |
| 4 | TB incidence 2020 | Burden | Calibration | 197 | 26 | 21 | 33.5 | as above |
| 5 | Active TB prevalence 2020 | Burden | Calibration | 304 | 10 | 34.7 | 36.1 | National TB Prevalence Survey 2019-21 |
| 6 | TB mortality 2020 | Burden | Calibration | 33 | 4.2 | 4.3 | 6.0 | WHO Global TB Report |
| 7 | NoLoop-only fraction | Cascade | Benchmark | 0.20 | 0.04 | 0.045 | 0.06 | Subbaraman et al. 2016 |
| 8 | Test-loop-only fraction | Cascade | Benchmark | 0.50 | 0.07 | 0.071 | 0.10 | Subbaraman et al. 2016 |
| 9 | Test+Tx-fail fraction | Cascade | Benchmark | 0.30 | 0.05 | 0.049 | 0.07 | Subbaraman et al. 2016 |
| 10 | Tests per tested episode, overall | Cascade | Benchmark | 3.5 | 0.55 | 0.55 | 0.78 | Cazabon et al. 2017; Subbaraman et al. 2016 |
| 11 | Tests per tested episode, no loop | Cascade | Benchmark | 1.0 | 0.30 | 0.29 | 0.42 | as above |
| 12 | Tests per tested episode, diagnostic loop only | Cascade | Benchmark | 3.0 | 0.65 | 0.65 | 0.92 | as above |
| 13 | Tests per tested episode, any treatment failure | Cascade | Benchmark | 4.5 | 0.90 | 0.91 | 1.28 | as above |

Note: Burden rates are per 100,000 population; cascade fractions are proportions, and testing statistics are diagnostic tests per episode that reached testing, the denominator the published benchmark uses; episodes that never reached a test are outside it. σ_total combines observational and model-discrepancy uncertainty; emulator variance is added separately during screening (Section S-3.3). Values are rounded.

#### S-3.3 Uncertainty specification and discrepancy decomposition

Table S-1 reports observational uncertainty (σ_obs), model-discrepancy uncertainty (σ_disc), and their combined uncertainty (σ_total). Observational uncertainty is estimated from published 95% confidence intervals, where available, by dividing the interval half-width by 1.96. Model discrepancy allows for model simplifications and differences between the empirical target and the quantity represented by the model. These components are combined as:

$$\sigma^2_{\mathrm{total},i} = \sigma^2_{\mathrm{obs},i} + \sigma^2_{\mathrm{disc},i}$$

For the four incidence targets, σ_total is approximately 17% of each point estimate, with discrepancy allowances of 21–34 per 100,000. The prevalence target is the survey's crude estimate of microbiologically confirmed pulmonary TB among people aged ≥15 years (304 per 100,000; 95% CI 285–324), rather than its adjusted estimate of 316 per 100,000. The confidence interval gives σ_obs ≈ 10 per 100,000. A discrepancy allowance of 34.7 per 100,000 was specified before calibration to accommodate differences from the model's all-age active TB prevalence and its single-node representation without explicit geography, giving σ_total = 36.1 per 100,000. Discrepancy allowances for mortality and the care-cascade benchmarks are reported in Table S-1.

For the six burden targets, emulator prediction variance is added to this combined variance when calculating implausibility. The resulting denominator is: $\sqrt{\sigma^2_{\mathrm{total},i} + \mathrm{Var}_{\mathrm{emul}}[f_i(x)]}$ The seven care-cascade benchmarks do not enter emulator screening or NROY construction. Their discrepancies are assessed from direct simulation outputs at the selected parameter set using σ_total alone.

#### S-3.4 Free parameters and priors

The ten free parameters describe transmission, TB natural history, care-seeking delays, and historical improvements in care and transmission (Table S-2). The secular transmission parameter reduces node infectivity geometrically from 1997 to 2024, after which it remains constant. It represents changes in transmission not explicitly captured by the care cascade; its prior includes zero, allowing no additional secular decline. The nominal untreated duration (3.5 years) and case-fatality fraction (0.55) used to derive natural cure and mortality rates were fixed before joint calibration. The resulting duration and fatality differ by disease presentation (Section S-1.2 and Table S-9).

Parameter choices and prior bounds were informed by published literature: care-seeking durations from Sreeramareddy et al. (2014)[10]; GeneXpert sensitivity from Cazabon et al. (2017)[9] and Steingart et al. (2014)[11]; pre-treatment fraction from Subbaraman et al. (2016)[8]; reactivation rate from Horsburgh (2010)[12]; re-infection susceptibility from Andrews et al. (2012)[13]; fast-progressor fraction from Vynnycky and Fine (1997)[14]; Base_Infectivity prior extended to encompass higher-transmission Indian settings reported in Mandal & Arinaminpathy (2015)[15] and Vesga et al. (2019)[6]. Parameters with strongly skewed plausible ranges (Base_Infectivity, Slow_Progressor_Rate, two care-seeking durations) use log-uniform priors; remaining parameters use uniform priors over the linear scale.

Table S-2. Free parameters varied during joint calibration

| # | Parameter (yr or d) | EMOD configuration | Min | Max | Scale | Selected value | Source |
|---|---|---|---|---|---|---|---|

| | | | | | | | |
|---|---|---|---|---|---|---|---|
| 1 | Base infectivity (per day) | Base_Infectivity_Constant | 0.005 | 0.100 | log | 0.0420 | Calibrated |
| 2 | Presymptomatic duration (yr) | TB_Presymptomatic_Rate | 0.15 | 0.50 | linear | 0.233 | Ku 2021[16]; Emery 2023[17] |
| 3 | Care-seeking delay, fast (d) | HSB probability_per_step | 10 | 30 | log | 14.2 | Calibrated |
| 4 | Care-seeking delay, slow (d) | HSB probability_per_step | 60 | 180 | log | 158 | Calibrated |
| 5 | Reactivation rate (per day) | TB_Slow_Progressor_Rate | 5e-6 | 5e-5 | log | 8.27e-06 | Calibrated |
| 6 | Re-infection susceptibility | Post_Infection_Acquisition_Multiplier | 0.20 | 0.80 | linear | 0.320 | Andrews 2012[13] |
| 7 | Fast-progressor fraction | TB_Fast_Progressor_Fraction_Adult | 0.05 | 0.20 | linear | 0.0765 | Calibrated |
| 8 | Annual gain in treatment initiation, 1997-2024 | DiagnosticTreatNeg / RampDTN treatment_fraction | 0 | 0.012 | linear | 0.00172 | Calibrated |
| 9 | Annual relative gain in care-seeking, 1997-2024 | HSB probability_per_step | 0 | 0.050 | linear | 0.01650 | Calibrated |
| 10 | Annual secular decline in transmission, 1997-2024 | NodeInfectivityMult | 0 | 0.020 | linear | 0.01283 | Calibrated |

Note: The ten parameters were calibrated together as combinations of values. The “Selected value” column lists the ten parameter values in the single set chosen for the main projections. Other combinations also matched the calibration targets: Table S-7 shows how widely each parameter could vary among the combinations retained after calibration. The selected values therefore should not be read as uniquely determined estimates. Diagnostic sensitivity was held fixed during calibration, whereas the historical increase in treatment initiation was calibrated. Section S-1.4 describes how the interventions change this calibrated baseline from 2025: Dx improves diagnostic sensitivity; Tx improves treatment initiation and cure.

#### S-3.5 Emulator specification

A separate Gaussian-process emulator with a Matérn-5/2 kernel is fitted to each calibration target using mogp_emulator [18], with scikit-learn’s GaussianProcessRegressor as a fallback [19]. The Matérn-5/2 kernel, commonly used in history matching[5][20], allows smooth responses without imposing the stronger smoothness assumption of the squared-exponential kernel [21]. Hyperparameters include a separate length-scale for each input dimension, the signal variance, and the noise variance. Separate length-scales capture differences in each target’s sensitivity to individual parameters, with larger length-scales indicating weaker sensitivity. Hyperparameters in mogp_emulator are estimated by maximum a posteriori optimisation.

The ten input parameters are scaled to [0,1] using their prior bounds, with parameters assigned log priors log-transformed before scaling. Burden outputs are also log10-transformed to stabilise variance, as their raw values span three orders of magnitude across the design and their variability increases with the mean. Each design point contributes the mean of its simulation replicates (10 in wave 1 and 25 in wave 2). The noise-variance term accounts for the Monte Carlo error remaining in these replicate means Scoring is carried out on the same log10 scale: the emulator moments are not back-transformed, and instead each target and its total uncertainty are transformed before scoring, the latter by the delta method as $\sigma/(z \ln 10)$. All six burden targets are treated this way, mortality included; Table S-1 reports the anchors on their original scale. [22].

#### S-3.6 Two-wave calibration design

Calibration used two waves: an initial exploration of the prior parameter space, followed by targeted sampling within the region not ruled out by wave 1. Wave 1 included 200 parameter combinations, each simulated 10 times (2,000 runs). Wave 2 added 100 combinations, each simulated 25 times (2,500 runs). The larger first wave provided broader coverage of the ten-dimensional input space, while the additional replicates in wave 2 reduced Monte Carlo noise. The 200+100 allocation followed the approach used by Vesga et al. [6]. For the initial design, 200 points also gave a lower space-filling discrepancy than 100 points (0.019 versus 0.035), indicating more even coverage.

Wave 1: exploring the prior space. We sampled 200 parameter combinations using a centred Latin hypercube with a fixed random seed and ran each under the baseline scenario. Emulators were fitted to log10-transformed outputs from simulations that passed an epidemic-viability screen, defined as 2020 incidence between 50 and 800 per 100,000. A random-forest classifier trained on all 200 combinations predicted whether unsimulated combinations would pass this screen. Log transformation was used because incidence varied across three orders of magnitude: initial fits on the raw scale were dominated by very high-incidence simulations and retained a much larger region than direct simulation results supported.

Wave 2: refining the plausible region. We sampled 100 additional combinations from the emulator-defined NROY region. Seventy were allocated to combinations representing a less effective present-day care cascade, defined by a 2024 treatment-initiation fraction below 0.82 and a care-seeking improvement multiplier below 2.0; the remaining 30 were sampled from the rest of the NROY region. This allocation increased coverage of the less effective cascade stratum while retaining combinations outside it; the latter need not be more effective on both measures.

Across both waves, 47 of 107 combinations in the less effective cascade stratum (43.9%) and 31 of 193 combinations in the other stratum (16.1%) were within 3σ of all six burden targets. Thus, the burden data remained compatible with a less effective present-day cascade. In wave 2 alone, 63 of 100 combinations passed this criterion. Parameter sets used in Section S-3.8 were selected from passing combinations in both strata.

The final emulators were fitted using eligible points from the combined 300-point design. Cross-validated $R^2$ ranged from 0.84 to 0.95, exceeding 0.90 for five of the six burden emulators. At the selected best fit, all six burden targets were reproduced within 0.3σ (maximum implausibility 0.27). The final emulator-defined NROY region occupied 5.76% of the prior volume. Calibration was stopped after two waves; the stopping rationale and unmet criteria are discussed in Section S-3.7.

Table S-3. Wave-by-wave summary of the calibration database

| Wave | Method | n design points | n valid | Max burden I at selected fit | Max benchmark I at selected fit | Retained fraction (denominator) |
|---|---|---|---|---|---|---|
| Wave 1 | Latin hypercube over the full 10-parameter prior | 200 | 78 | 1.24 | 1.20 | 7.5% (15/200) |
| Wave 2 | Emulator NROY, stratified 70/30 toward a loose 2024 cascade | 100 | 99 | 0.27 | 1.96 | 63% (63/100) |
| Combined | Final emulators fitted to 177 valid points from the combined design | 300 | 177 | 0.27 | 1.96 | 5.76% of the prior |

Note: A “valid” point passes the initial epidemic screen (2020 incidence of 50–800 per 100,000). The retained fractions of 7.5% (15/200) and 63% (63/100) are the proportions of simulated parameter combinations meeting all six burden targets within the allowed uncertainty. The final 5.76% instead estimates the share of the entire prior parameter space retained by the combined emulators. These percentages have different denominators and cannot measure how much the retained region shrank between waves. At each selected parameter set, “Max burden I” is the largest discrepancy among the six burden targets; “Max benchmark I” is the largest among the seven cascade benchmarks. The benchmark score is evaluated at the set selected for its burden fit. Table S-6, rows 1–2, repeats these scores to compare them with the threshold of 3.

#### S-3.7 Calibration stopping criteria and benchmark comparison

Table S-4 summarises the criteria used to assess calibration adequacy and guide stopping. The six burden targets were used to identify the not-ruled-out-yet (NROY) region, whereas the seven care-cascade benchmarks were assessed only at the selected best fit and did not influence NROY construction. The benchmarks therefore provide a check on care-cascade features that were not directly targeted during calibration.

Table S-4. Calibration assessment and stopping criteria

| # | Criterion | Metric | Threshold | Source | Action if violated |
|---|---|---|---|---|---|
| 1 | Implausibility at the best fit (calibration targets) | min over x of max over the six burden targets of I_i(x) | ≤ 3.0 (3σ rule) | Andrianakis 2015[22]; Vesga 2019 | Reconsider the σ_disc allocation; add a wave 3 |
| 2 | Implausibility at the best fit (benchmarks) | max over the seven cascade targets of I_i at the best fit | All 7 within 3σ | Andrianakis 2015[22] | Re-examine structural assumptions rather than re-fitting |
| 3 | Emulator validity | 5-fold cross-validated $R^2$ of each burden emulator (log10 scale) | $R^2 > 0.9$ | Convention adopted in this study | Increase the design size; reconsider the kernel |
| 4 | NROY volume (diagnostic) | V_NROY,wave2 / V_prior | Reported; < 5% expected | Andrianakis 2015[22] (informativeness benchmark) | If still large, add a wave 3 (50 design points within the residual NROY) |
| 5 | Wave-to-wave NROY reduction (diagnostic) | V_NROY,wave2 / V_NROY,wave1 | < 50% | Convention adopted in this study | If contraction is limited, assess whether another wave would add information |
| 6 | Cross-wave KL divergence (diagnostic) | D_KL ( π_NROY, wave2 ‖ π_NROY, wave1 ) | < 0.1 nats | Convention adopted in this study | If divergence is large, assess whether further refinement is needed |

Note: This table defines the assessment criteria; Table S-6 reports the results. Criteria 1–3 assess agreement with burden data, agreement with cascade benchmarks, and emulator accuracy. Criterion 4 describes the size of the retained parameter region; its final value is reported in Table S-6, row 4. Criteria 5–6 assess how much that region shrinks and how its parameter distribution changes between waves. These two measures were not calculated after the final emulators were refitted using both waves, so their thresholds were not demonstrated to have been met. Rows 5*–6* of Table S-6 are separate incidence checks, not results for criteria 5–6 here.

Calibration was stopped after two waves. At the selected best fit, the maximum implausibility across the six burden targets was 0.27, well below the threshold of 3. All seven care-cascade benchmarks were also within 3σ in the full-run cohort evaluated during calibration (maximum 1.96; Table S-6). In the 2015–2024 episode cohort used for the main-text trajectory analyses, six benchmarks remained within 3σ, while the no-loop fraction differed by 3.2σ (Table S-5). The model assigned more episodes to the no-loop group than the published cascade implied. This difference was interpreted as a structural mismatch between the published sequential cascade stages and the longitudinal episode groups used here, rather than as a target for further fitting. The test-loop-only fraction (0.26 against 0.50; Table S-5, row 8) is mostly a consequence of mutual exclusivity: 49% of episodes with a diagnostic loop later failed treatment and are counted in the treatment-failure group, and the share of episodes with any diagnostic loop is 0.51.

Table S-5. Per-target implausibility at the calibrated best fit, production run

| # | Target | z [σ_total] | Production run | I_i | ≤ 3σ? |
|---|---|---|---|---|---|
| 1 | TB incidence 2000 | 322 [54.7] | 313.7 | 0.15 | Yes |
| 2 | TB incidence 2010 | 276 [46.9] | 262.1 | 0.30 | Yes |
| 3 | TB incidence 2015 | 237 [40.3] | 228.6 | 0.21 | Yes |
| 4 | TB incidence 2020 | 197 [33.5] | 197.6 | 0.02 | Yes |
| 5 | Active TB prevalence 2020 | 304 [36.1] | 291.0 | 0.36 | Yes |

| | | | | | |
|---|---|---|---|---|---|
| 6 | TB mortality 2020 | 33 [6.0] | 30.7 | 0.38 | Yes |
| 7 | NoLoop-only fraction | 0.20 [0.06] | 0.393 | 3.21 | No (3.2) |
| 8 | Test-loop-only fraction | 0.50 [0.10] | 0.262 | 2.38 | Yes |
| 9 | Test+Tx-fail fraction | 0.30 [0.07] | 0.345 | 0.65 | Yes |
| 10 | Tests per tested episode, overall | 3.5 [0.78] | 2.64 | 1.10 | Yes |
| 11 | Tests per tested episode, no loop | 1.0 [0.42] | 1.00 | 0.00 | Yes |
| 12 | Tests per tested episode, diagnostic loop only | 3.0 [0.92] | 2.38 | 0.67 | Yes |
| 13 | Tests per tested episode, any treatment failure | 4.5 [1.28] | 4.16 | 0.26 | Yes |

Note: Direct-simulation implausibility is |model − z|/σ_total. Results use 100 eligible replicates. Rows 1–6 are burden targets; rows 7–13 are cascade benchmarks for episodes activating in 2015–2024. Treatment-failure-only episodes are included in the treatment-failure group. Row 8 counts diagnostic loops only in episodes without treatment failure; the share of episodes with any diagnostic loop is 0.51. Values are rounded.

Not all assessment criteria were met. Cross-validated $R^2$ exceeded 0.90 for five of the six burden emulators, but was 0.84 for mortality. The selected parameter set nevertheless reproduced the mortality target within 0.4σ. The final NROY region occupied 5.76% of the prior volume, slightly above the 5% diagnostic benchmark. Given the close fit to the burden targets and the full-run cascade benchmarks, no third wave was run; whether further refinement would change the selected parameter set remains untested.

For direct comparisons at the selected parameter set, both burden targets and care-cascade benchmarks use the absolute model–data difference divided by σ_total, with a threshold of 3. Emulator screening additionally includes prediction variance and applies only to the six burden targets. A close fit at the selected set therefore does not by itself establish emulator accuracy throughout the retained region.

Table S-6. Calibration assessment results and additional incidence checks

| # | Criterion | Threshold | Wave 1 | Wave 2 / final assessment | Type |
|---|---|---|---|---|---|
| 1 | Max implausibility over calibration targets | ≤ 3 | 1.24 | 0.27 | Required |
| 2 | Max implausibility over benchmarks | ≤ 3 | 1.20 | 1.96 | Required at the best fit (3.2σ on the production cohort; S-3.7) |
| 3 | Emulator cross-validated $R^2$ (burden) | > 0.9 | 0.79-0.88 | 0.84-0.95 (5 of 6 targets > 0.9) | Required (partially met) |
| 4 | Retained fraction (different denominators) | Reported | 7.5% of design | 5.76% of prior | Reported |
| A | Held-out incidence years (2005, 2023) | Within 1σ of the WHO estimate | not assessed | 2005: 264.7 vs 311 ± 52.9 (0.88σ); 2023: 190.0 vs 192 ± 32.6 (0.06σ) per 100,000 | Reported |
| B | Implied 2000-2020 incidence trend | Compared with WHO -2.43%/yr | -2.17%/yr | -2.28%/yr (production) | Reported |

Note: Rows 1–3 report results for the first three criteria in Table S-4. Row 2 assesses cascade benchmarks across the full simulation period. The largest benchmark discrepancy is therefore 1.96 here, whereas the no-loop fraction differs by 3.2σ in that later cohort. Row 4 reports the final retained parameter-space volume for criterion 4 (5.76% of the prior); its wave-1 value (7.5%) is a pass rate among simulated points and cannot be compared directly with that volume. Table S-5 instead reports individual benchmark results for TB episodes activating in 2015–2024 in the production simulations. Rows A, B are additional incidence checks; Row A compares predictions with incidence data for 2005 and 2023, neither used for calibration. Row B summarises the decline between the 2000 and 2020 calibration years and is not an independent validation.

The seven cascade metrics—three composition fractions and four tests-per-case statistics—are used as benchmarks to assess care pathways without directly fitting their composition. These metrics depend strongly on lower-quality diagnostic sensitivity and the treatment-initiation fraction, which are modified in the Dx and Tx scenarios, respectively.

Lower-quality diagnostic sensitivity is held fixed during calibration. The historical trend in treatment initiation is calibrated, but the Tx scenario specifies improvement relative to the resulting 2024 value.

#### S-3.8 Parameter uncertainty and its implications for projections

Calibration narrowed the combinations of parameters compatible with the burden targets more than it narrowed individual values. Only base infectivity and the slow-progression rate were narrowed appreciably (retained spans of 52% and 64% of the prior); the other eight free parameters kept 87–90% (Table S-7). This is expected: history matching discards parameter sets more than 3σ from any target without ranking those that remain, and six aggregate burden targets cannot fix ten parameters separately. The retained region is a ridge along which base infectivity trades off against progression. Prior draws are independent, so any correlation among retained draws is induced by the targets: infectivity is negatively correlated with the slow-progression rate (Spearman ρ = −0.68), the fast-progressor fraction (−0.35) and the reinfection multiplier (−0.30), and within terciles of infectivity the median slow-progression rate falls from 0.55 to 0.18 of its prior range. The burden targets thus constrain how many infections occur and how quickly they become disease, not either factor alone. The operational parameters (care-seeking delays and the three long-term trends) are uncorrelated with every other parameter, with retained medians within 0.03 of the prior centre: the same incidence trajectory can be produced by several mixtures of earlier treatment, earlier care-seeking and declining transmission, and these quantities genuinely differ across states, providers and periods.

The care-cascade benchmarks could not resolve this. Cascade composition is set mainly by inputs held fixed in the design, such as test sensitivity and the baseline treatment-initiation fraction; among the 78 simulated design points meeting the 3σ criterion on all burden targets, each benchmark varied by only 0.1–0.5 of its tolerance (for example, the test-loop-only fraction 0.26–0.35 against ±0.10), so including the benchmarks as targets would not have excluded any of these points. The benchmarks do respond to the operational parameters within this narrow range (tests per case with the slow-subgroup delay, ρ = −0.82; the test-loop-only fraction with the treatment-initiation gain, −0.90), but the published values are too uncertain, and in part defined differently from the model groups (note to Table S-5), to discriminate among parameter sets. The same stability has a positive side: the loop-group composition on which the main-text burden decomposition rests varies by only about 0.03 across burden-consistent parameter sets, so that decomposition, like the intervention effects in Table S-8, is largely insensitive to the calibration uncertainty described here.

The selected parameter set is therefore one point on this ridge rather than a uniquely identified estimate, and values within the marginal intervals of Table S-7 cannot be combined freely. Intervals in the main text describe stochastic variation at the selected set; the twenty-set propagation below samples along the ridge, so the across-set intervals in Table S-8 reflect the infectivity–progression trade-off and the unconstrained operational parameters. Estimating individual parameters would require targets that respond to one factor but not the other, such as the subclinical fraction for the natural-history parameters and time-to-treatment distributions for care-seeking, or a likelihood-based step within the retained region; further waves of history matching alone cannot supply it.

Table S-7. Marginal extent of the NROY region for each free parameter

| Parameter | Prior range | NROY 5-95% | Retained span | Production value |
|---|---|---|---|---|
| Base infectivity (per day) | 0.005 - 0.1 | 0.01676 - 0.08076 | 52% | 0.04196 |
| Presymptomatic duration (years) | 0.15 - 0.5 | 0.1661 - 0.4804 | 90% | 0.233 |
| Care-seeking delay, fast subgroup (days) | 10 - 30 | 10.61 - 28.35 | 90% | 14.17 |
| Care-seeking delay, slow subgroup (days) | 60 - 180 | 63.86 - 171.6 | 90% | 157.8 |
| Reactivation rate, slow progressors (per day) | 5.00e-6 - 5.00e-5 | 5.49e-6 - 2.42e-5 | 64% | 8.27e-6 |
| Reinfection susceptibility multiplier | 0.2 - 0.8 | 0.2151 - 0.7359 | 87% | 0.3197 |
| Fast-progressor fraction, adults | 0.05 - 0.2 | 0.05603 - 0.1906 | 90% | 0.07647 |
| Treatment-initiation gain (per year) | 0 - 0.012 | 5.33e-4 - 0.01114 | 88% | 0.00172 |

| Parameter | Prior range | NROY 5-95% | Retained span | Production value |
|---|---|---|---|---|
| Care-seeking improvement (per year) | 0 - 0.05 | 0.002753 - 0.04727 | 89% | 0.0165 |
| Secular infectivity decline (per year) | 0 - 0.02 | 0.001014 - 0.01879 | 89% | 0.01283 |

Note: Intervals are marginal 5th–95th percentiles of retained points from 400,000 prior draws after viability and implausibility screening. Retained span is measured on the prior sampling scale (log scale for log-uniform parameters). These describe NROY geometry, not posterior credible intervals; an unchanged uniform prior would have a 90% central span.

To assess sensitivity to parameter choice, we selected 20 sets from the 80 simulated combinations that met the 3σ criterion for all six burden targets. Selection was stratified by the secular-decline parameter and included the set used for the main projections. Each set was simulated under all eight scenarios with 10 replicates, giving 1,600 runs. Intervention effects were calculated relative to each set's own baseline and summarised across sets using the median and 2.5th–97.5th percentiles (Table S-8).

Table S-8. Projected reductions in 2035 (%) at the selected parameter set and across 20 retained sets

| **Scenario vs 2015** | **Incidence Selected set** | **Incidence Across sets** | **Prevalence Selected set** | **Prevalence Across sets** | **Mortality Selected set** | **Mortality Across sets** |
|---|---|---|---|---|---|---|
| Baseline | 35.1 [31-38] | 38.7 [18-56] | 37.9 [35-40] | 43.5 [25-58] | 41.4 [33-49] | 45.9 [29-72] |
| ACF | 45.8 [43-49] | 49.5 [31-60] | 56.8 [55-59] | 60.8 [47-69] | 64.5 [59-70] | 68.6 [49-82] |
| Dx | 36.9 [34-40] | 42.8 [19-57] | 41.8 [39-44] | 46.8 [29-61] | 42.9 [35-50] | 48.6 [21-67] |
| Tx | 46.8 [44-50] | 49.4 [37-61] | 69.2 [68-71] | 72.3 [66-78] | 65.7 [61-70] | 67.8 [44-84] |
| ACF+Dx | 48.8 [46-52] | 51.9 [38-63] | 60.6 [59-62] | 61.6 [52-70] | 69.7 [65-74] | 68.7 [58-82] |
| ACF+Tx | 55.2 [53-58] | 55.6 [44-66] | 84.7 [84-86] | 83.3 [76-88] | 85.6 [82-88] | 85.3 [73-94] |
| Dx+Tx | 47.8 [44-51] | 52.0 [34-62] | 72.0 [71-73] | 73.7 [65-78] | 67.3 [61-73] | 73.8 [50-83] |
| ACF+Dx+Tx | 56.9 [54-59] | 57.3 [45-68] | 85.4 [85-86] | 83.7 [77-88] | 88.5 [86-91] | 87.3 [76-94] |
| **Scenario vs 2035 baseline** | **Incidence Selected set** | **Incidence Across sets** | **Prevalence Selected set** | **Prevalence Across sets** | **Mortality Selected set** | **Mortality Across sets** |
| ACF | 16.5 [13-20] | 13.3 [7-33] | 30.5 [28-33] | 26.2 [16-45] | 39.4 [29-49] | 36.3 [5-63] |
| Dx | 2.7 [-1 to 6] | 3.6 [-4 to 15] | 6.3 [3-10] | 6.2 [-7 to 11] | 2.6 [-12 to 16] | 3.4 [-95 to 45] |
| Tx | 18.0 [15-21] | 16.0 [6-35] | 50.4 [48-52] | 51.3 [42-60] | 41.5 [33-49] | 37.7 [-6 to 71] |
| ACF+Dx | 21.1 [18-24] | 18.7 [8-40] | 36.6 [34-39] | 32.6 [17-51] | 48.3 [40-56] | 44.2 [-35 to 62] |
| ACF+Tx | 31.0 [28-34] | 24.1 [14-46] | 75.3 [74-76] | 69.5 [59-80] | 75.4 [70-80] | 73.3 [32-88] |
| Dx+Tx | 19.5 [16-22] | 16.9 [7-37] | 54.9 [53-57] | 54.4 [46-62] | 44.2 [33-54] | 47.7 [9-63] |
| ACF+Dx+Tx | 33.6 [31-36] | 28.9 [13-50] | 76.4 [75-77] | 71.0 [57-81] | 80.4 [76-84] | 75.9 [63-86] |

Note: Selected-set estimates use 100 eligible replicates and bootstrap 95% CIs; across-set estimates show medians [2.5th–97.5th percentiles] from 20 sets with 10 replicates each. Reductions use ratios of replicate means. Negative values indicate increases relative to the comparator; across-set intervals are not confidence or posterior credible intervals.

The main qualitative findings persisted across the parameter sensitivity analysis, although effect sizes varied (Table S-8). Diagnostic improvement alone had a small median incidence effect, with an across-set interval that included no reduction. Treatment support produced a larger median prevalence reduction than active case finding, whereas their median incidence reductions were similar. The combined package produced much larger reductions in prevalence and mortality than in incidence; its reported incidence interval relative to 2015 remained well below the 90% target. These results support the conclusions within the parameter sets examined, while leaving uncertainty in fixed parameters and values outside the prior bounds untested.

#### Text S-4. Model parameter listing

Table S-9 lists the principal demographic, natural-history, HIV, diagnostic, and treatment parameters. Jointly calibrated values correspond to the selected set in Table S-2; other entries are fixed assumptions or derived quantities, as indicated.

Table S-9. Model parameters, fixed assumptions, and derived quantities

| Parameter | Value | Unit | Description | Source |
|---|---|---|---|---|
| *Demographic, temporal, and geographic-mixing parameters.* | | | | |
| Base year | 1850 | year | Simulation day 0 | Model architecture (spin-up start) |
| burn_initial | 730 | days | Initial burn-in (2 yr) | Model architecture |
| burn_predots | 52,925 | days | Pre-DOTS burn-in (145 yr) | Model architecture (1852-1997 pre-DOTS) |
| To_end_from_DOTS | 14,965 | days | Legacy constant; the production run sets Simulation_Duration directly so that the run ends in 2050 | Model architecture (1850-2050) |
| x_Base_Population | 20 | multiplier | Population-size scaling | Computational scaling (this study) |
| Base_Individual_Sample_Rate | 0.05 | — | Within-population sampling rate | EMOD computational sampling |
| x_Birth | 0.75 | multiplier | Birth-rate scaling | Calibrated to UN WPP 2019 (India CBR)[23] |
| QualityOfCare High / Low | 15% / 85% | — | Population share in higher- vs lower-quality care streams | Subbaraman et al. 2016[8]; Pai et al. 2016[24] |
| Geographic HINT groups | Low / Mid / High (33/34/33%) | — | Spatial heterogeneity | Modelling assumption; India NTPS 2019-21 regional variation |
| HINT within-group BI | 3.1 / 2.9 / 3.5 | — | Within-group infectivity | Calibrated to NTPS regional variation (this study)[25] |
| HINT between-group BI | 0.1 | — | Between-group infectivity | Modelling assumption (low cross-region mixing) |
| ART start year | 2003 | year | Antiretroviral therapy rollout | NACO India ART rollout records[26] |
| HIV epidemic start | 1982 | year | ~15 yr before DOTS | UNAIDS India epidemic estimates[27] |
| *Disease natural-history parameters.* | | | | |
| Base_Infectivity_Constant | 0.0420 | per day | Per-day infectivity (calibrated) | Calibrated (this study, wave-2 design 4) |
| Post_Infection_Acquisition_Multiplier | 0.320 | — | Re-infection susceptibility | Andrews 2012[13]; McCreesh & White 2018[7] |
| TB_Fast_Progressor_Fraction_Adult | 0.0765 | — | Fast-progressor fraction (adults) | Vynnycky & Fine 1997[14]; Horsburgh 2010[12] |
| TB_Fast_Progressor_Fraction_Child | 0.03 | — | Fast-progressor fraction (children) | Marais et al. 2004[28] |
| TB_Fast_Progressor_Rate | 0.0175 | per day | Fast-progression rate | EMOD-TBHIV default; Vynnycky & Fine 1997[14] |
| TB_Slow_Progressor_Rate | 8.27e-06 | per day | Reactivation rate (~0.3% / yr) | Calibrated (this study); cf. Horsburgh 2010[12] |
| TB_Presymptomatic_Rate | 0.01176 | per day | Transition to symptomatic | Calibrated derivative (this study) |
| TB_Presymptomatic_Duration_Years | 0.2330 | years | Presymptomatic phase (calibrated) | Calibrated (this study); Drain et al. 2018[29]; Kendall & Dowdy 2025[30] |
| TB_Active_Presymptomatic_Infectivity_ Multiplier | 0.166 | — | Presymptomatic infectiousness | Fixed design value (this study); relative to smear-positive disease. |
| TB_Smear_Positive_Fraction_Adult | 0.65 | — | Smear-positive fraction (adults) | India NTPS 2019-21; Steingart et al. 2006[31] |
| TB_Smear_Positive_Fraction_Child | 0.25 | — | Smear-positive fraction (children) | Marais et al. 2004 |

| Parameter | Value | Unit | Description | Source |
|---|---|---|---|---|
| TB_Smear_Negative_Infectivity_Multiplier | 0.346 | — | Smear-negative infectiousness | Behr et al. 1999[32]; [32] Tostmann et al. 2008 [33] The value of 0.346 sits above the 0.24 (95% CI 0.20-0.30) reported by Tostmann et al. and is a deliberately conservative allowance for transmission from smear-negative disease. |
| TB_Extrapulmonary_Fraction_Adult | 0.10 | — | Extrapulmonary fraction (adults) | India NTPS 2019-21; NTEP Annual Report[34] |
| TB_Extrapulmonary_Fraction_Child | 0.40 | — | Extrapulmonary fraction (children) | Marais et al. 2004 |
| TB_Extrapulmonary_Mortality_Multiplier | 0.15 | — | Extrapulmonary mortality multiplier | EMOD-TBHIV default; Tiemersma et al. 2011[35] |
| TB Duration (untreated) | 3.5 | years | Nominal mean untreated duration used to derive the cure and mortality rates; realised 3.6 yr across presentations after the x1.5 mortality multiplier | Fixed (this study); Tiemersma et al. 2011[34] |
| Death Fraction | 0.55 | — | Nominal untreated case fatality ratio; realised 0.65 for smear-positive disease and 0.53 across presentations after the x1.5 mortality multiplier | Fixed design value (this study); Tiemersma et al. 2011.[34] Presentation-specific fatality follows from the derived rates and mortality multipliers. |
| TB_Active_Cure_Rate | $3.52\times10^{-4}$ | per day | Natural cure rate, derived as (1 - death fraction) / duration | Derived from TB Duration 3.5 yr and Death Fraction 0.55 (this study); Tiemersma 2011[34] |
| TB_Active_Mortality_Rate | $6.458\times10^{-4}$ | per day | Mortality rate derived from the nominal case-fatality fraction and cure rate, with a ×1.5 multiplier. Implied smear-positive duration ≈2.75 years and case fatality ≈0.65. | Calibrated derivative; Tiemersma 2011[35] |
| TB_Immune_Loss_Fraction | 0 | — | Post-cure immunity loss | Modelling assumption (no waning) |
| TB_Relapsed_to_Active_Rate | 0.012 | per day | Relapse-to-active rate | Panjabi et al. 2007[36] |
| TB_MDR_Fitness_Multiplier | 1.0 | — | MDR fitness cost | Modelling assumption (DR not modelled) |
| TB_Enable_Exogenous | 1 | — | Exogenous reinfection enabled | McCreesh & White 2018[7] (re-infection enabled) |
| *HIV epidemiology and TB co-infection dynamics.* | | | | |
| HIV epidemic start | 1982 | year | ~15 yr before DOTS | UNAIDS India epidemic estimates |
| ART start | 2003 | year | Antiretroviral therapy | NACO India ART rollout records |
| ART_Reactivation_Factor | 1.0 | — | TB reactivation on ART | Modelling baseline; Suthar et al. 2012[37] |
| CoInfection_Mortality_Rate_Off_ART | $3.0\times10^{-4}$ | per day | TB/HIV mortality (no ART) | Williams et al. 2010[38]; Suthar et al. 2012[37] |
| CoInfection_Mortality_Rate_On_ART | $1.0\times10^{-4}$ | per day | TB/HIV mortality (on ART) | Williams et al. 2010[38]; Suthar et al. 2012[37] |
| TB_CD4_Activation_Vector | $[5\times10^{-4}, 2.5\times10^{-4}, 1\times10^{-4}, 5\times10^{-5}, 2.8\times10^{-5}]$ | per day | CD4-stratified activation | EMOD-HIV default; Williams et al. 2010[38] |
| HIV prevalence in adults (peak 2005) | 0.5% | — | Indian peak prevalence | UNAIDS India peak prevalence ~2005[39] |
| ART coverage by 2020 | 0.80 | — | WHO ART coverage estimate | WHO/UNAIDS India 2020 |
| *Diagnostic-cascade parameters by historical era.* | | | | |
| Care-seeking rate (fast, calibrated) | 1 / 14.2 | per day | ~14-day delay in 1997, ~9 d by 2025 (calibrated improvement) | Calibrated (this study); Sreeramareddy et al. 2014[10] |
| Care-seeking rate (slow, calibrated) | 1 / 158 | per day | ~158-day delay in 1997, ~101 d by 2025 (calibrated improvement) | Calibrated (this study); Sreeramareddy et al. 2014[10] |

| Parameter | Value | Unit | Description | Source |
|---|---|---|---|---|
| Care-seeking budget | 7 | visits | Max attempts per episode | Modelling assumption; Kapoor et al. 2012 [40] |
| Pre-DOTS Sensitivity (High QoC) | 0.70 / 0.70 | SmPos/SmNeg | Public sector smear | Steingart et al. 2006[31] |
| Pre-DOTS Sensitivity (Low QoC) | 0.60 / 0.60 | SmPos/SmNeg | Private sector smear | Cazabon et al. 2017[9] |
| Pre-DOTS Treatment fraction | 0.60 | — | Pre-DOTS Tx initiation | Subbaraman et al. 2016[8] |
| DOTS Treatment fraction | 0.70 | — | DOTS-era Tx initiation | India NTEP / RNTCP records |
| GeneXpert Sensitivity (High QoC) | 0.75 / 0.65 | SmPos/SmNeg | GeneXpert, with operational and specimen losses; the pre-DOTS smear sensitivities (row above) are optimistic placeholders for an era with little documented performance data, which is why this value is lower | Steingart et al. 2014 (Cochrane)[11] |
| GeneXpert Sensitivity (Low QoC) | 0.65 / 0.60 | SmPos/SmNeg | Smear + clinical dx | params.py (this study); Steingart et al. 2006[31] |
| GeneXpert-era treatment fraction (both QoC) | 0.70 in 1997 -> 0.746 in 2024 | — | Calibrated secular ramp, not a fixed value; the treatment-support scenario halves the residual loss relative to the 2024 value | Calibrated (this study); Subbaraman et al. 2016[8] |
| GeneXpert introduction year | 2012 | year | CBNAAT rollout | India NTEP CBNAAT rollout records |
| GeneXpert ramp-up duration | 8 | years | Scale-up 2012-2020 | India NTEP scale-up records (2012-2020) |
| Mean re-test interval after false-negative | 60 | days | ~8 weeks | Fixed modelling assumption |
| Mean re-engagement after pre-Tx default | 60 | days | Same as false-negative | Modelling assumption |
| *Treatment-pathway parameters.* | | | | |
| Standard regimen duration | 180 | days | Drug-sensitive regimen | WHO 2010 TB Treatment Guidelines[41] |
| Regimen cure proportion | 0.80 (DOTSHQ) / 0.50 (DOTSLQ) | — | Probability of cure per completed course, by care stream | emodpy-tbhiv standard drug definitions; Subbaraman et al. 2016[8] |
| Mean delay before re-treatment after failure | 180 | days | ~6 months until restart | Modelling assumption (NTEP follow-up cycle) |
| Infectious window during treatment | 14 | days | First ~2 weeks before sterilisation | Jindani et al. 2003[42] |
| Treatment-failure relapse multiplier | 1.0 | — | Relative infectiousness of relapse | Modelling assumption |
| Drug-drop probability per failed course | 0.0 | — | No additional drop modelled | Modelling assumption (second-line not modelled) |
| NodeInfectivityMult (secular decline) | 0.01283 | per yr | Annual geometric decline in node infectivity 1997-2024 (x0.71 by 2024); proxy for non-cascade drivers of the historical decline | Calibrated (this study) |
| Treatment-initiation gain | 0.00172 | per yr | Annual absolute gain in the treatment-initiation fraction, applied equally to both care streams, 1997-2024 (0.70 -> 0.746) | Calibrated (this study) |
| Care-seeking gain | 0.01650 | per yr | Annual relative gain in care-seeking probability, 1997-2024 (x1.56 by 2024) | Calibrated (this study) |
| Low-quality stream cure proportion | 0.50; 0.80 from 2025 under Tx | - | From 2025, Tx raises the lower-quality stream cure proportion from 0.50 to 0.80 using a separate regimen, preserving historical treatment outcomes. | emodpy-tbhiv standard drugs |

#### Text S-5. Proposed trajectory-sensitive, transmission-weighted indicators for TB care-cascade monitoring

Table S-10. Proposed indicators of repeated care, delays, and transmission-relevant burden in TB care pathways

| Indicator | Definition | What it adds | Conventional measure complemented |
| --- | --- | --- | --- |
| **Diagnostic-loop index** | Mean number of diagnostic testing encounters before treatment initiation per confirmed case, distinguishing repeat encounters after negative results or incomplete care from scheduled or confirmatory testing. | Identifies repeated diagnostic encounters that may prolong time without treatment. | Test positivity; diagnostic yield |
| **Treatment-loop index** | Mean number of treatment episodes, including restarts, per treated patient. | Identifies repeated treatment following interruption, failure, or recurrence. | Treatment success rate |
| **First-contact-to-cure interval** | Median days from first healthcare contact to documented durable cure, among patients achieving cure. | Captures the overall duration of the care pathway, including delays and repeated episodes. This is not equivalent to infectious duration. | Time to treatment initiation |
| **Pre-cascade infectious time** | Estimated duration of infectious disease before first diagnostic contact, including subclinical disease and patient delay. | Captures infectious time occurring before patients enter diagnostic care. | No direct equivalent |
| **Low-loop completion rate** | Percentage of successfully treated patients who reached cure without diagnostic or treatment loops. | Distinguishes success through a single-pass pathway from success after repeated care episodes. | Treatment success rate |
| **Transmission-adjusted cascade** | Cascade performance weighted by estimated infectious person-time averted relative to a specified comparator, accounting for relative infectiousness. | Connects care improvements to their potential contribution to reducing transmission. | Cascade coverage and retention |
| **Retest interval** | Median days from a negative test or an incomplete diagnostic encounter to the next diagnostic test within the same episode. | Shows whether repeat testing occurs promptly, even when the number of tests does not change. | Tests per case |

Note: Indicators require consistent definitions and linked patient histories; measures of unobserved infectious time also require modelling. Report unresolved pathways separately. The transmission-adjusted measure requires a defined comparator and further validation.

The proposed indicators shift attention from whether patients complete care to how they reach that outcome and how much infectious time accumulates along the way. Similar treatment-success rates can conceal repeated failures and prolonged delays, while unchanged testing counts can mask improvements from faster follow-up. These measures therefore complement conventional cascade indicators by distinguishing fewer failures from faster resolution of failures. The indicators are intended as complementary monitoring concepts and require validation before routine use.
